# Obsessive-compulsive disorder secondary to focal brain lesions: from lesions to networks

**DOI:** 10.1101/2025.01.09.25320060

**Authors:** Gonçalo Cotovio, Nelson Descalço, Jaime Caballero-Insaurriaga, Daniel Martins, Francisco Faro Viana, Catarina Fonseca, João Ramos, Ana Maia, José Oliveira, Nuno Loução, Shan H. Siddiqi, Michael D. Fox, J. Bernardo Barahona-Corrêa, Albino J. Oliveira-Maia

## Abstract

**Background:** Obsessive-compulsive disorder (OCD) may develop following brain lesions, but lesion distribution and connectivity patterns are unknown.

**Methods:** OCD-associated lesions, identified from systematic literature search, were traced on common brain space and compared to control lesions (N=608). Topography was analyzed using brain atlases, and lesion location networks computed using normative functional connectivity (N=1000). Lesional network maps (LNMs) were contrasted to data from primary OCD.

**Results:** Among 129 case-descriptions of lesional OCD, traced lesions (n=40) were more specifically located in orbitofrontal cortex bilaterally and right temporal pole, and more connected to orbitofrontal cortex and ventral basal ganglia, bilaterally. LNMs overlapped with primary OCD functional peaks from Neurosynth, revealed abnormal functional connectivity in patients with primary OCD (n=54) compared to controls (n=61), and aligned with brain stimulation targets.

**Conclusions:** Lesional OCD has specific patterns of brain lesion topography and functional connectivity, with LNMs associated to brain functional patterns in primary OCD.

## INTRODUCTION

Obsessive-Compulsive Disorder (OCD) is a potentially incapacitating psychiatric disorder with an estimated lifetime prevalence of 1.3%^1^. Although the exact cause cannot be identified in most cases, OCD is sometimes diagnosed *de novo* after focal brain insults of different etiologies^2–5^. The location of such injuries varies across existing reports^6^, raising questions whether brain lesions causing OCD-like symptoms follow a particular distribution in the brain, damaging specific regions or circuits.

Exploring the neuroanatomy of lesional neuropsychiatric disorders can help establish causal links between brain regions or circuits and specific syndromes^7^, while potentially revealing new treatment targets for therapeutic brain stimulation strategies^8,9^. Several neuroimaging analysis methods, namely pooled lesion topography analysis^8^ and lesion network mapping^9,10^, can be used to systematically define neuroanatomic models of lesional neuropsychiatric syndromes. Lesion topography analysis assesses the anatomical overlap of different lesions across anatomically-defined brain regions. Lesion network mapping, on the other hand, assesses the overlap of lesion location connectivity, to estimate impact on functionally connected brain networks, leveraging normative resting state functional connectivity data, obtained from large databases of healthy individuals^11,12^. Both strategies have been used successfully to define dysfunctional brain regions and circuits across several lesional neuropsychiatric syndromes^8–10^. While cases of lesional obsessive-compulsive syndromes have been reported in the literature^2–5^, pooled lesion topography analysis and lesion network mapping for lesional OCD is not yet available.

In this study we tested the hypothesis that focal brain lesions associated with the development of obsessive-compulsive disorder follow a preferential anatomical and functional network distribution. Furthermore, we aimed to explore the overlap of anatomical and/or functional patterns identified for lesional OCD with functional patterns found in primary OCD, to address the possibility that knowledge from brain lesions may be relevant to enhance understanding of mechanisms underlying non-lesional OCD, with potential application in the development of novel treatments for this disorder.

## MATERIALS AND METHODS

### Lesional OCD Cohort

Cases of lesional OCD were obtained from literature search according to a predefined protocol, following PRISMA guidelines^13^. The protocol was published in PROSPERO database (CRD42020200330) and can be consulted for full description of methods^14^. Search was performed on PubMed, Web-of-Science, EMBASE and PsycINFO. Articles in English, Portuguese, Spanish, German, or French were considered, irrespective of publication year or country of origin. The full list of Search Terms (Table S1) as well as the Inclusion and Exclusion Criteria applied to this study is available in *Supplementary Material*. Two investigators independently assessed eligibility and extracted clinical and socio-demographic data, with consensus obtained, if necessary, consulting at least one senior psychiatrist (JO, JBB-C, and/or AJO-M). For each eligible case, extracted data included age at onset of OCD symptoms, sex, hand dominance, time-interval between brain insult and first OCD-like symptoms, symptom domains (e.g. obsessions, compulsions, hoarding), availability of an anatomical representation of the brain lesion, authors’ description of lesion location, brain lesion etiology, patient’s personal and family history of other neuropsychiatric disorders, medication at OCD symptoms onset, medication used to treat OCD symptoms, duration of symptoms, length of follow-up, recurrence of symptoms, and study quality according to previously published quality assessment tools – Clinical Quality Assessment scale (CQA) and Brain Lesion Documentation Assessment scale (BLDA)^8^. When anatomical representation of brain lesions was available, a team of two researchers (CF and JR), one of whom is a neuroradiologist, jointly traced eligible lesions manually (i.e. BLDA≥3) onto a standardized brain atlas (Montreal Neurological Institute; MNI)^15^ using MITK software^16^. Only tissue damage that was clearly visible in the available images was traced, according to the different orientation planes provided, and no extrapolation to juxtaposed brain slices was performed (two-dimensional images; 2D). A third researcher (GC), with expertise in lesion tracing, then independently reviewed each lesion tracing.

### Control Lesions Cohort

A cohort of control lesions (N=608), which was considered as an approximation to the normative anatomical distribution of brain lesions, was defined using openly available lesions that have not been selected for any particular symptom presentation, namely the BRATS dataset (N=369) for brain tumor lesions^17–19^ and the ATLAS R 1.2 dataset (N=239) for stroke lesions^20^. Contrary to lesional OCD cases, that were 2D, the control lesions cohort consisted of three-dimensional (3D) brain scans. To ensure comparability across the two cohorts, and according to previously published methodology, we sampled only some brain scan slices per patient in the control lesions cohort^8^. Briefly, in a Python 3 notebook, for each scan in the control lesions cohort, we found the slice with maximum damage extent and considered it the main slice. Additional neighboring lesion-including slices, at 8 slices intervals above and/or below the main slice, were obtained for some control lesions, according to the number of slices available per lesion in the lesional OCD cohort, for matching purposes. The distribution of slices in the control sample in relation to orientation plane (x, y, z) was also defined according to the corresponding proportion in the lesional OCD cohort, rendering the two groups comparable.

### Lesional OCD topography

The overlap of OCD-associated lesions on a 3D space for voxel-based lesion-symptom mapping^21^ is underestimated when using 2D lesion tracings. Thus, quantification of lesion distribution in grey-matter (GM) and white-matter (WM) was performed using lesion location analysis, as described previously^8^. This was performed in a Python 3 notebook using the Automated Anatomical Labeling atlas (AAL)^22^ for GM, and the John Hopkins University (JHU) WM tractography atlas^23^ for WM. For each area of the GM and WM atlases, we calculated the proportion of affected voxels, and performed the more conservative one-sample Sign Test, where symmetry assumption is not required, to find the areas intersected by OCD lesions, and compute the “Lesional OCD Topography Map”. We considered one-sided p-values, as negative proportion of affected voxels within each atlas area is not possible in this condition. We then compared the proportion of affected voxels in each area of the GM and WM atlases, with that observed in the control lesions cohort (N=608) using general linear models, controlling for lesion size and lesion etiology (vascular vs. non-vascular), to produce the “Lesional OCD vs. Controls Topography Map”. Peak regions of these maps were considered if false discovery rate (FDR) corrected p-value (p_FDR_) <0.05. Please see *Statistical Analysis* section for further details. In an exploratory analysis, within the lesional OCD cohort we further compared, for each area of AAL and JHU atlases, the proportion of affected voxels on the left-vs. right-hemisphere, using the sign test for each AAL and JHU region^8^ and considering two-sided p-values, as negative values are possible in this analysis.

### Lesional OCD functional connectivity

In order to obtain a functional connectivity map of lesional OCD, we computed the connectivity map of each lesion location as described previously in other studies using *lesion network analysis*^9^. Succinctly, we used resting state functional magnetic resonance imaging (rs-fMRI) data from a large human dataset (N=1000)^11^, commonly known as normative functional connectome, to perform this analysis. Each OCD or control lesion was used as a region of interest (ROI) to perform seed-based connectivity across the processed human connectome data^24,25^. Within each subject of the connectome, the average rs-fMRI time series of a lesion was extracted and then correlated with the timeseries of each voxel, resulting in a map of Pearson’s correlation coefficients (r). Fisher transformation was applied to the map and maps were then averaged across all subjects of the human connectome, generating a unique mean normative connectivity map per lesion location, i.e., a lesion connectivity map performed for each lesion of the lesional OCD and control lesion cohorts. A “Lesional OCD Mean Connectivity Map” was obtained from the connectivity T-map for each OCD lesion, calculated using a one-sample T-Test as described previously^25–30^, and then averaged across all OCD lesions. The “Lesional OCD vs. Controls Connectivity Map” was obtained from the original per-lesion connectivity maps, that were compared between OCD and control lesions, identifying brain regions that are more specifically connected to lesions causing OCD-like symptoms (i.e., a specificity map)^9^. Peak regions of this map were considered when family-wise error (FWE) corrected p-value (p_FWE_) <4.1×10^−5^ in a volume ≥200 mm^3^. This threshold was selected through visual inspection, by consensus between a group of researchers (GC, ND, JBB-C and AJO-M), overlaying the map on a standard brain and adjusting the threshold to guarantee a balance between the cluster size and their grey matter overlap, aiming to minimize partial volume effects^31^. Please see *Statistical Analysis* section for further details. Finally, a binary “Intersection Map” was obtained as the intersection of the Lesional OCD vs. Controls Connectivity Map peak regions, with a thresholded version of the Lesional OCD Mean Connectivity Map. The threshold was again defined by consensus between a group of researchers (GC, ND, JBB-C and AJO-M) to maximize connectivity overlap to at least one brain region while increasing region definition^25–27^ (T>±6.5, i.e., p_FWE_<1.4×10^−5^).

### Testing reliability and robustness of topography and connectivity maps

Both reliability and robustness of topography and connectivity maps were assessed using Pearson’s spatial correlation between the maps obtained in the various sensitivity analyses, and the maps that resulted from the main analyses. To test the reliability of the process for manually tracing of individual lesions, we assessed if lesion topography and connectivity maps where replicable when lesions were traced by a different researcher, independent of the originally tracing team (inter-rater reliability), or by the original tracing team in two separate sessions (intra-rater reliability). We analyzed intra- and inter-rater reliability between lesion tracings for size and localization, as well as with new Lesional OCD vs. Controls topography and connectivity maps. To assess reliability of methods for analysis, we repeated computation of the Lesional OCD vs. Controls Topography Map using an alternative statistical approach, as performed previously^8^. For connectivity methods, we repeated computation of connectivity maps using a different rs-fMRI human brain connectome^12^ (N=937), as done previously^32^ (please see *Supplementary Material* for additional preprocessing steps that were required^33^ for *lesion network analysis*). Additionally, to test if the Lesional OCD vs. Controls Connectivity Map was being disproportionately driven by lesions located over the connectivity peak differences, i.e., auto-connectivity, we re-computed the map after excluding all brain lesions that overlapped the connectivity peak regions obtained in the original analyses. In data-driven assessment of reliability, we randomly divided each cohort in two equally sized subgroups (N_OCD_: 20 vs. 20; N_Controls_: 304 vs. 304), to produce two entirely separate datasets for analysis.

To assess robustness, Lesional OCD vs. Controls Topography and Connectivity Maps were re-computed in several sensitivity analyses, restricting inclusion of OCD cases to assess the impact of potential confounders, specifically: fulfilment of strict DSM 5 criteria for OCD; moderate to severe OCD symptom severity according to Y-BOCS total score; short latency between lesion occurrence and symptom onset according to median split (symptom onset ≤240 days after lesion); no concomitant medication; no personal or family history of psychiatric disorders; only insults causing well circumscribed focal brain lesions (i.e., excluding cases with traumatic or infectious etiology); high quality brain imaging documentation (BLDA≥4); MRI scans; high quality clinical reporting (CQA≥4). Although we computed Lesional OCD vs. Controls Topography and Connectivity Maps while controlling for lesion etiology, the nature of control lesions^34^ did not match all the etiologies comprised in the lesional OCD cohort. Hence, we also repeated computation of both Lesional OCD vs. Controls Maps, excluding tumor controls, excluding vascular controls, excluding non-vascular and non-tumor OCD lesions, and restricting both OCD and control cohorts only to vascular etiology. Restricting the two cohorts to tumoral lesion cases yielded an excessively low sample size in the lesional OCD arm (N=7), that did not allow for that sub-analysis.

### Convergence of lesional network maps with primary OCD data

We proceeded to assess if functional connectivity networks emerging from analyses of lesional OCD are relevant to understand the neurobiology of primary idiopathic OCD. We first tested the overlap between the Lesional OCD vs. Controls Connectivity Map with functional ROIs associated on meta-analysis with symptoms of OCD, and compared the result with the overlap with functional ROIs associated with other neuropsychiatric syndromes. The functional ROIs for primary OCD and other neuropsychiatric syndromes were generated from Neurosynth^35^, a tool that meta-analyzes neuroimaging data that are associated with a given search term^36^. In addition to “OCD”, we searched for the terms “Aphasia”, “Anxiety”, “Cognitive Deficits” and “Depression”, because these were the most prevalent neuropsychiatric co-morbidities in our lesional OCD cohort (please see Results section for more details). From Neurosynth, we obtained a binary map of the voxels most associated with each term, designated as functional ROIs. Overlap with our Lesional OCD vs. Controls Connectivity Map was obtained by computing the mean intensity value of voxels in that map within functional ROIs for each term. In a ROI-wise approach, to test if overlap was significantly different from what would be expected at chance level, the previous step was repeated for functional ROIs of each term with 10 000 random permutations of the lesional group label (OCD | Control), to obtain in each permutation a different Lesional OCD vs. Controls Connectivity Map (i.e., 10 000 permuted maps). Thus, for the functional ROIs of each term, we obtained a probability distribution of within-ROIs mean-intensity, used to test the null-hypothesis that the real within-ROIs mean-intensity was not significantly different than what is expected by chance. Additionally, in a voxel-wise approach, we tested, separately for voxels with positive and negative connectivity in the Lesional OCD vs. Controls Connectivity Map, if overlap of the voxels associated to “OCD” differed significantly from those in other syndromes, using Bonferroni-corrected two-sample t-tests.

In another approach, we used resting state functional resonance imaging (rs-fMRI) data from an observational case-control study ongoing in our lab (ClinicalTrials.gov reference NCT06566781). This study compared patients with primary, idiopathic OCD, diagnosed according to the Structured Clinical Interview for the Diagnostic and Statistical Manual of Mental Disorders 5^th^ Edition (SCID-5), with community-based healthy volunteers, frequency-matched by gender and age. Please see *Supplementary Material* for further details on eligibility criteria and neuroimaging data acquisition and preprocessing methods. Here, we explored if the functional connectivity networks associated to lesional OCD also differed between patients with primary OCD and healthy volunteers. Using a hypothesis-driven approach, we used a multivariate distance matrix regression (MDMR)^37–40^ to test if primary OCD patients and healthy subjects have a different functional connectivity pattern to a seed region. MDMR is a linear regression performed on a between-subjects dissimilarity matrix, in which each element represents a measure of dissimilarity (e.g., Euclidean distance) in high-dimensional data (e.g., functional connectivity maps) between a pair of subjects. The analysis thus tests whether the connectivity pattern is more similar within a group than between groups of subjects. In our case, MDMR analysis was run on the whole brain connectivity pattern to the region that reflects the intersection between the Lesional OCD Mean Connectivity and Lesional OCD vs. Controls Connectivity, i.e., the lesional connectivity nodes that are both most sensible and specific for the occurrence of OCD after a brain insult. First, for each subject of the primary OCD and healthy subjects’ samples, we computed a functional connectivity map to the aforementioned seed region, by calculating the Pearson correlation coefficient between the average time series of the seed region and those of each brain voxel. Then, we applied a Fisher z (Fz) transformation to the resulting coefficients, obtaining a final functional connectivity map for each individual. To compute the distance matrix, we subsampled the functional connectivity maps by a factor of 2 (4 mm isotropic), extracted the values from gray-matter voxels, and computed the Euclidean distance between all possible pair of subjects. We then run the MDMR analysis using an in-house Python 3 pipeline. In follow-up exploratory analyses, to identify specific brain regions that are differently connected to the lesional OCD connectivity intersection seed, we compared connectivity between the primary OCD cohort and healthy controls, for ROIs containing voxels with between-group mean connectivity differences exceeding 0.06 (Fz) in the same direction over a continuous volume greater than 120 mm³. Using an in-house Python 3 pipeline, after extracting mean Fz values in these ROIs for each individual, we performed region-wise Bonferroni corrected two-sample t-tests, to test if functional connectivity to the lesional intersection seed differed across patients with OCD and healthy volunteers.

Finally in an exploratory approach, we assessed whether the lesional OCD network map included target regions for invasive or non-invasive therapeutic neuromodulation for primary OCD. Specifically, we overlaid deep brain stimulation (DBS) and transcranial magnetic stimulation (TMS) targets reported in the literature for OCD treatment, with the Lesional OCD Mean Connectivity Map and the Lesional OCD vs. Controls Connectivity Map. For DBS we used four targets reported in the literature to be effective for OCD treatment: anteromedial subthalamic nucleus (amSTN)^41^; anterior limb of internal capsule (ALIC)^42,43^; ventral capsule/ventral striatum (VC-VS)^41^; and nucleus accumbens (Nac)^44^. We extracted the average MNI coordinates for these different targets from available literature: Left (−8, −14, −9) and Right (8, −13, −9) amSTN^41^; Left (−15.29, 8.08, 1.57) and Right (15.29, 8.08, 1.57) ALIC^45^; Left (−13, 8, −1) and Right (13, 8, −2) VC-VS^41^; Left (−3.78, 5.08, −7.79) and Right (3.78, 5.08, −7.79) Nac^45^. We created 3mm spherical ROIs for each of these DBS targets, approximating previous reports of DBS volume tissue activation^46^, centered at each coordinate. For TMS targets we considered the medial prefrontal cortex and anterior cingulate cortex (mPFC & ACC) as these have been the regions targeted by the TMS devices cleared and/or approved by international regulatory agencies for OCD treatment^47^. Centered at the MNI coordinate extracted from a previous report^48^ (0, 30, 30), we created a 10mm sphere ROI^49^.

### Statistical analysis

In *lesion topography analyses*, statistical testing was performed as described above and significance was defined according to Benjamini-Hochberg^50^, assuming a FDR of 0.1, to account for multiple comparisons^8^. In *lesion network analysis*, the comparison between OCD and control lesions’ connectivity was performed using voxel-wise permutation-based two-sample t-tests, implemented using FSL PALM^51^, two-tailed testing, threshold-free cluster enhancement (TFCE) and family-wise error (FWE) correction options, according to neuroimaging best practices^52^, controlling for lesion size and lesion etiology (vascular vs. non-vascular). We used permutation-based statistics followed by FWE correction, using 5000 permutations per test and α<0.05, decreasing the likelihood of false positive results^32,34,53^. To test if the connectivity profile of lesional OCD intersection nodes was different between primary OCD patients and healthy subjects, we used the MDMR analysis following the optimization described previously by others^40^. Briefly, we used a design matrix that contained the intercept and the clinical group (primary OCD patients vs. Healthy Subjects). The MDMR pseudo-F statistic was extracted for the group effect and P-value extracted after permuting the data 10 000 times. We repeated this analysis controlling for age, sex, and dose of antidepressant medication, expressed as the equivalent daily dose of fluoxetine in mg^54^. As medication and group were highly collinear, medication was orthogonalized to group^55^. Missing values were handled by imposing the within-group mean. All statistical analyses were performed in StataCorp. 2017. Stata Statistical Software: Release 15. College Station, TX: StataCorp LLC, unless mentioned otherwise.

## RESULTS

### Lesional OCD cohort characterization

From an initial literature search of 3592 articles, 73 articles, published between 1982 and 2020, including 129 case-descriptions, were eligible (Figure 1). Obsessions and compulsions were the most frequently reported manifestations (94.4%) and were similarly frequent (76.4% and 80.9%, respectively). Since other obsessive-compulsive spectrum disorders, namely body dysmorphism (0.8%), hoarding (5.4%), trichotillomania (2.3%), or excoriation (1.6%), were seldomly described, the term OCD will be used for simplicity. Mean age at lesional OCD onset was 46.4±16.9 years old and most patients were male (59.6%), contrary to what has been described in primary OCD (32.3%^56^), and right-handed (91.3%). A prior history of a neuropsychiatric diagnosis prior to the first OCD symptoms was reported in 41.9%, epilepsy being the most common. The average time between the brain insult and the first OCD symptoms was just over three months (i.e., 97.1 days), and the most frequent lesional etiology was vascular (27.9%). On average, OCD symptoms were reported to resolve after 5 years, with the follow-up period after OCD-like symptom-onset varying between 1 month and 44 years. Co-occurring neuropsychiatric symptoms/syndromes associated with brain insult occurred in 62.0% of cases, including cognitive deficits (50.4%), depressive symptoms (31.0%), change in personality traits (24.0%), aphasia (17.1%) and anxiety symptoms (10.9%). At the onset of OCD only 12.4% of patients were reported to be taking any type of medication, with antihypertensives being the most common (2.3% of all patients). Only 5.4% were taking antiepileptics, antidepressants, antipsychotics, benzodiazepines and/or other central nervous system agents. Medication to alleviate lesional OCD symptoms was reported in 33.3% of cases, with selective serotonin reuptake inhibitors being the most frequent (72.1%), and only 15.5% of all cases were reported to receive psychological therapy/support.

**Figure 1.**
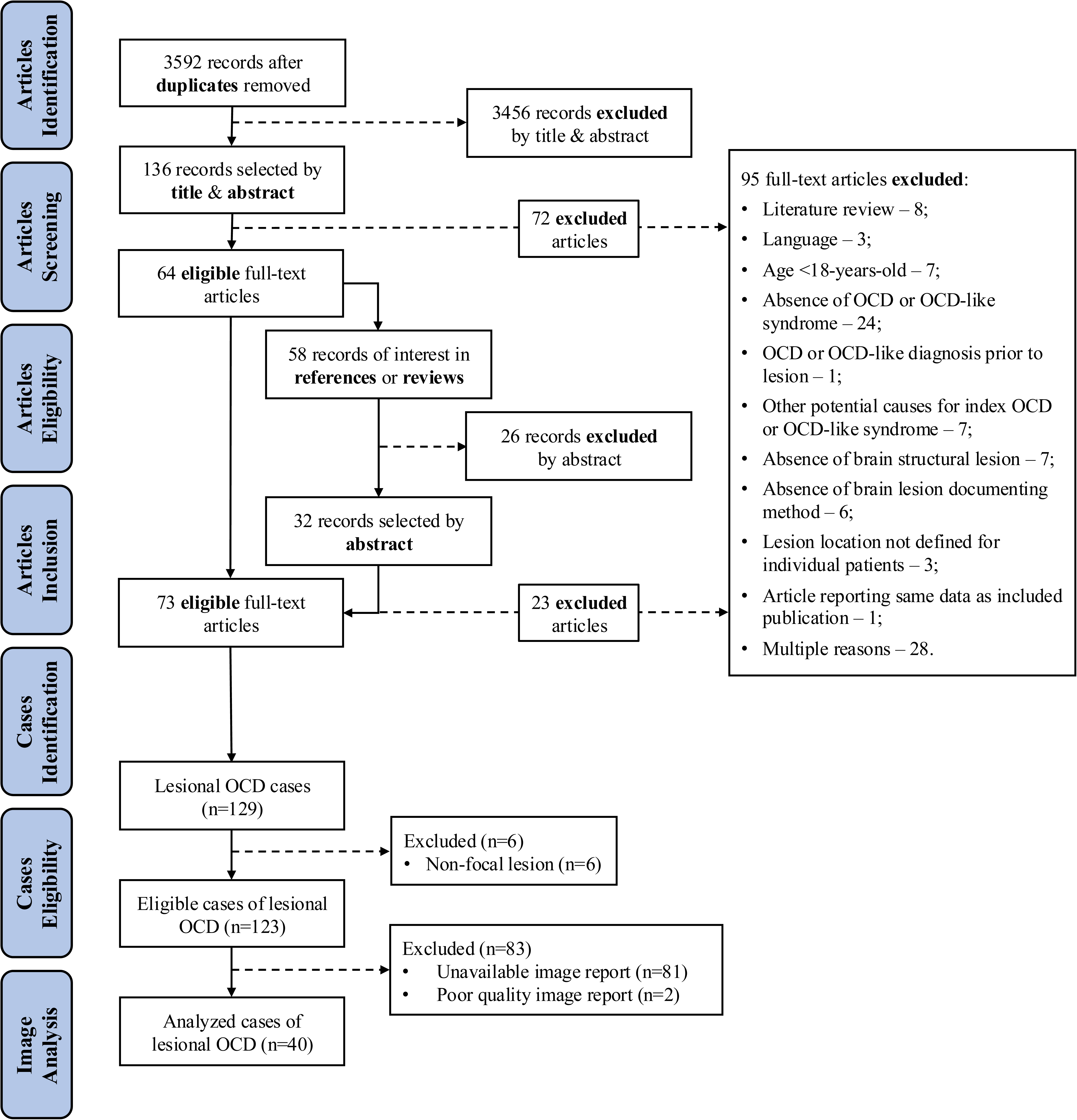
PRISMA flowchart. From 73 eligible articles included in the systematic review we extracted 123 cases with focal lesions, 40 of which were eligible to be traced. OCD – obsessive compulsive disorder; PRISMA – preferred reporting items for systematic reviews and meta-analyses.

### Lesional OCD topography

From the 129 case-descriptions, 123 cases were associated with focal brain lesions, i.e., involving at least one circumscribed brain area^57^. Focal lesions mostly affected the temporal lobes (48.0%), basal ganglia (30.1%) and frontal lobes (28.5%), with similar distribution between right and left hemispheres (p>0.08, McNemar’s test; Figure 2A). From these 123 cases, images of brain lesion were provided for 42 patients, 2 of which were excluded due to poor image quality (BLDA<3; Figure 1), and the remaining 40 traced on the MNI atlas (Figure 1). This subset was representative of the full sample, with only 2 cases (5%) with an OCD spectrum disorder rather than OCD, and traced lesions also occurring more frequently in the temporal lobe, basal ganglia, and frontal lobe (Figure 2B), and equally in both hemispheres (Table S2). When compared to control lesions (Figure S1A), OCD lesions were more frequently located in the orbitofrontal cortex (OFC), bilaterally (p_FDR_<0.004, for several regions of the OFC; Figure 2C; Table S3), and in the right temporal pole (p_FDR_<0.005; Figure 2C; Table S3).

**Figure 2.**
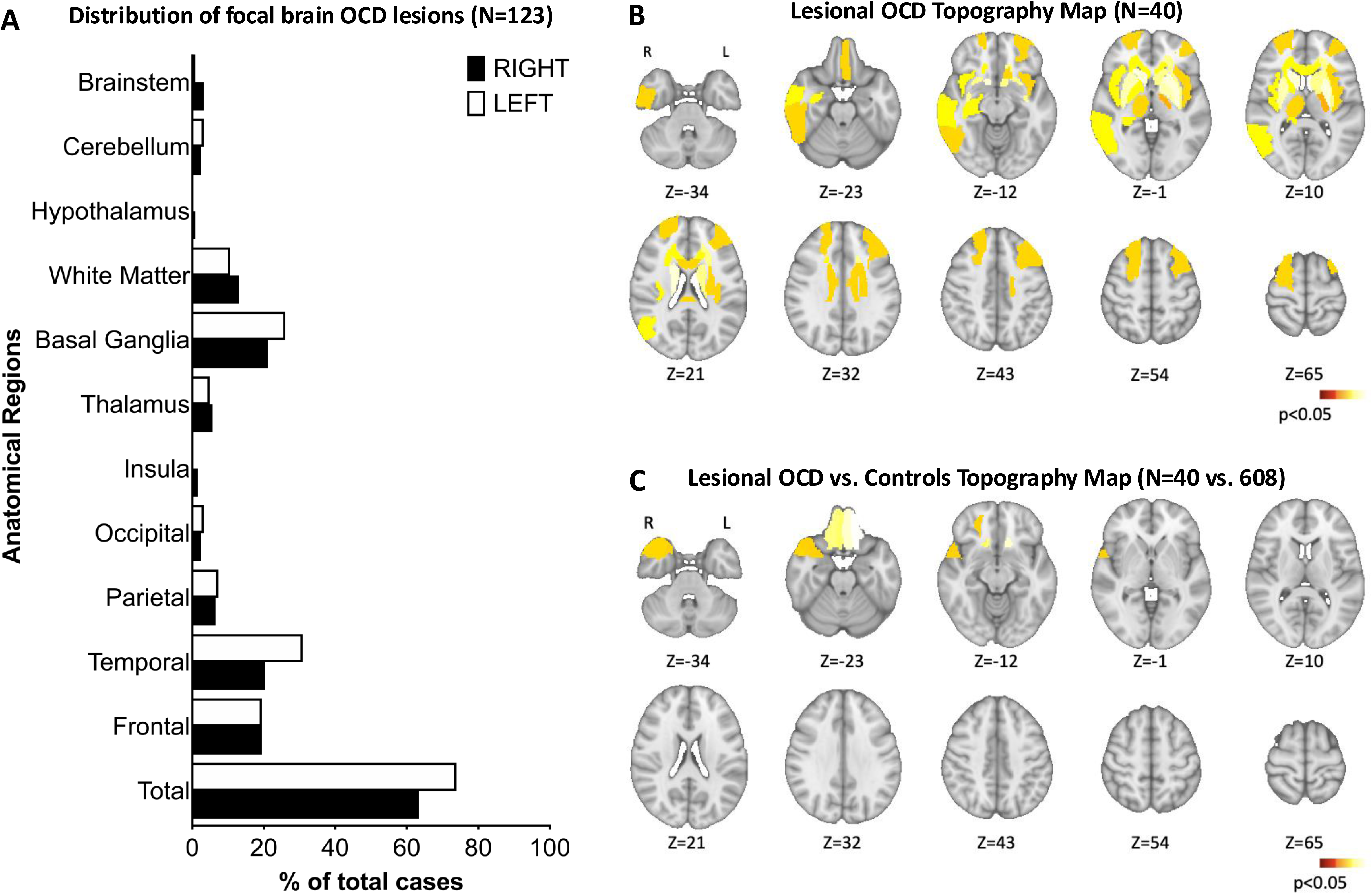
Topography of OCD lesions. **A.** We identified 123 cases with focal brain lesions causing OCD or OCD spectrum symptoms. Description of lesions revealed preferential locations in temporal lobes (48.0%), basal ganglia (30.1%) and frontal lobes (28.5%) with no significant differences between right and left hemispheres (p>0.05 for all, McNemar’s test). B. Similarly, lesion images (N=40) revealed significant lesion location overlap in the temporal lobes, basal ganglia and frontal lobes, also with no significant differences between right and left hemispheres (p>0.05 for all, sign test; Table S2). C. When compared to control lesions (N=608), lesions causing OCD were more frequently located bilaterally in the OFC and in the right middle and superior temporal pole. Maps in panels B and C were corrected for multiple comparisons according to Benjamini-Hochberg^50^ and are displayed at an FDR-corrected level of p<0.05. Warm colors show regions with higher likelihood for occurrence of lesions associated with OCD. FDR – false discovery rate; OCD – obsessive compulsive disorder; OFC – orbitofrontal cortex.

### Lesional OCD functional connectivity

Normative functional connectivity of OCD lesions was computed using *lesion network mapping*^9^, as detailed above. Briefly, for each lesion location we computed the mean connectivity with every brain voxel, using resting-state fMRI data from a large sample of healthy volunteers, publicly available in a human brain connectome^24,25^ (Figure S1B). Mean functional connectivity across lesion locations resulted in a Lesional OCD Mean Connectivity Map. To identify brain regions specifically connected to OCD lesion locations, we statistically compared the mean connectivity of control lesion locations to the Lesional OCD Mean Connectivity Map, thus obtaining a specificity map of OCD lesional connectivity^9^, designated as the Lesional OCD vs. Controls Connectivity Map (Figure S1B). OCD-associated lesions had widespread normative connectivity, converging preferentially onto the basal ganglia, bilaterally (Figure 3A). However, when comparing with connectivity of control lesions, the OFC and ventral basal ganglia had significantly higher connectivity to OCD lesions, bilaterally (Figure 3B). Peak regions in the Lesional OCD vs. Controls Connectivity (p_FWE_<4.1×10^−5^) were distributed in the frontal poles, medial and orbital frontal cortices, accumbens, caudate and putamen, bilaterally (Figure S2), and intersected, in part, a thresholded (T>±6.5; p_FWE_<1.4×10^−5^) version of the Lesional OCD Mean Connectivity Map (Figure 3C).

**Figure 3.**
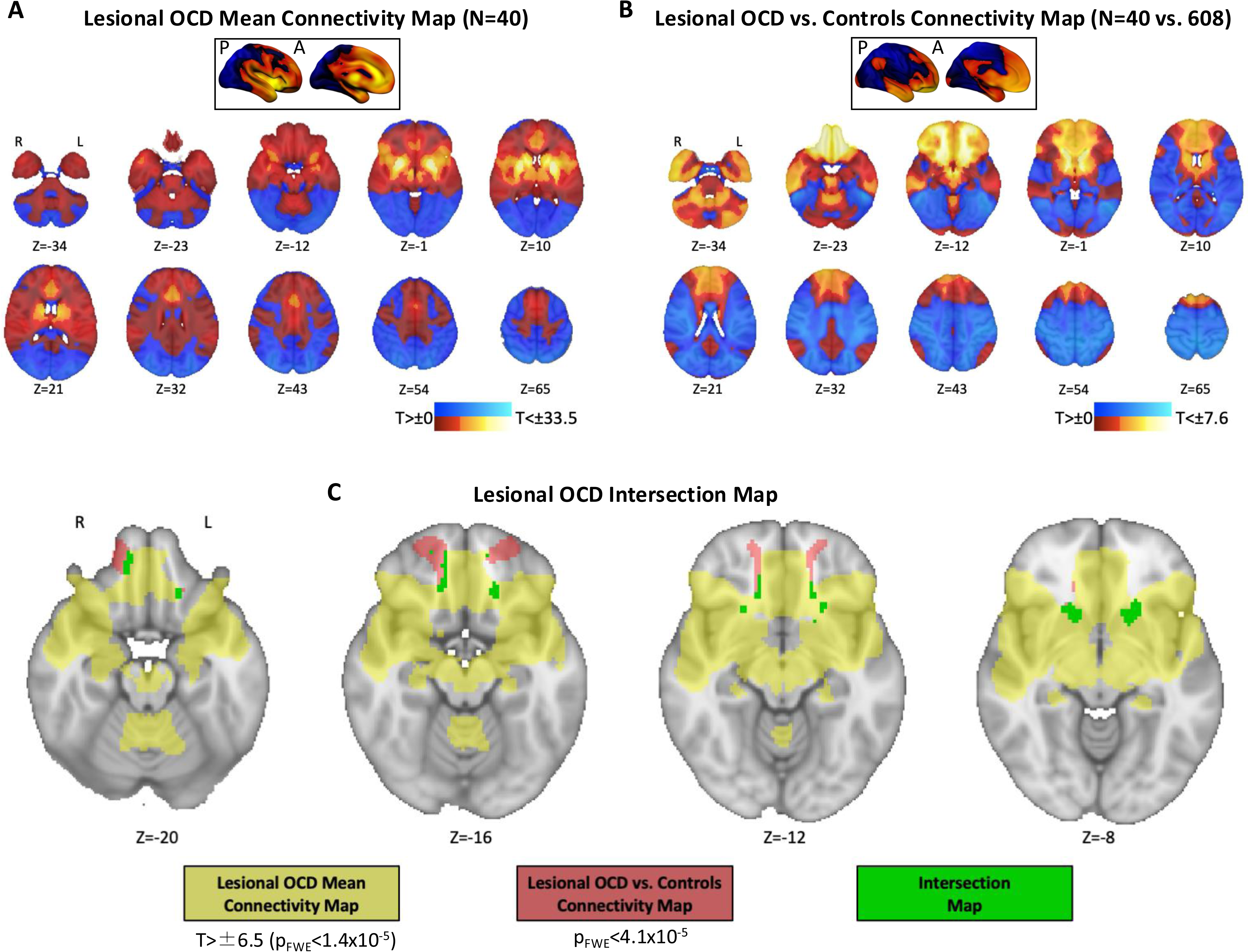
Connectivity of OCD lesions. **A.** The Lesional OCD Mean Connectivity Map reflects mean connectivity of OCD lesion locations (N=40), that was distributed across many cortical and subcortical regions, with preferential location in the basal ganglia, bilaterally. Warm and cold colors represent areas that are more positively or negatively connected to OCD lesions, respectively. **B.** For the Lesional OCD vs. Controls Connectivity Map, OCD lesion connectivity was compared to control lesions (N=608), showing that lesions causing OCD were more connected to the OFC and ventral basal ganglia, bilaterally. Warm and cold colors represent areas that are more or less connected to OCD lesions when compared to controls, respectively. **C.** Regions of peak connectivity, obtained when intersecting thresholded maps for Lesional OCD Mean Connectivity (T>±6.5; p_FWE_<1.4×10^−5^) and Lesional OCD vs. Controls Connectivity (p_FWE_<4.1×10^−5^), were distributed in the frontal pole, frontal medial and orbital cortices and the ventral basal ganglia, bilaterally. FWE – family wise error; OCD – obsessive compulsive disorder; OFC – orbitofrontal cortex.

### Reliability and robustness of lesional OCD topography and network maps

In line with previous work in lesional mapping^8,9,58^, we focused on the Lesional OCD vs. Controls Maps, since those are specific to lesional OCD, rather than representing specificities of lesion etiology (e.g., vascularization), and tested if they were methodologically reliable and robust to potential confounders. Topography and connectivity maps were significantly overlapping with original maps, after replacing the original OCD tracings by those performed by an independent tracer or by the original tracing team in a different occasion (inter-rater or intra-rater reliability; r≥0.94, spatial correlations with original maps; Table S4). Reliability of the topography map was supported by analyses using distinct statistical methods (r=0.76), while that of the connectivity map was supported by use of a distinct connectome (r=0.85) as well as analyses excluding lesions overlapping peak connectivity regions, to address auto-connectivity (r>0.9; Table S4, Figure S3). Finally, results were also consistent when OCD and control data was randomly split into two-halves to repeat analyses in separate datasets (r>0.81; Table S4). Robustness of the two maps was assessed in several sensitivity analyses, restricting inclusion criteria according to clinical characteristics related to OCD (diagnostic criteria, symptom severity, medication at symptom onset, neuropsychiatric risk factors; r>0.78; Table S4) or the brain lesion (lesion to symptom latency, lesion etiology; r>0.86; Table S4), and according to quality of reporting or images (r>0.93; Table S4). Since the nature of control lesions did not encompass all the lesion etiologies present in the lesional OCD cohort, we further explored the impact of lesion etiology in Lesional OCD vs. Controls Maps. We found that regions identified in the topography map differed when restricting OCD and/or control lesions to specific etiologies (Figure S3A), while connectivity peak regions remained consistent (Figure S3B).

### Convergence of functional connectivity in lesional and primary OCD

We investigated if our lesion-based results are relevant to known pathophysiology of primary, idiopathic OCD. To address this question, we first tested how functional ROIs associated with different neuropsychiatric syndromes (Figure 4A-E), as obtained from the meta-analytic tool Neurosynth, overlapped with the OCD vs. Controls Connectivity Map. Overlap with the functional ROIs occurred above chance-level for “OCD” (Figure 4A; p<0.001), but not for symptoms/syndromes co-morbid with OCD in our lesional OCD sample, namely “Depression” (Figure 4B), “Anxiety” (Figure 4C), “Cognitive Deficits” (Figure 4D) and “Aphasia” (Figure 4E; p>0.05 for all, Bonferroni-corrected permutation tests; Figure 4F). In a voxel-wise approach, we confirmed that voxels in the OCD vs. Controls Connectivity Map with greater connectivity to OCD lesions were over-represented in Neurosynth “OCD” functional ROIs, relative to functional ROIs of other symptoms (p<0.001 for all). For voxels with greater connectivity to control lesions, a difference relative to “OCD” functional ROIs was found only for the “Cognitive Deficits” functional ROIs (p<0.001), but not for other symptoms (p>0.05 for all, Bonferroni-corrected t-tests; Figure 4G).

**Figure 4.**
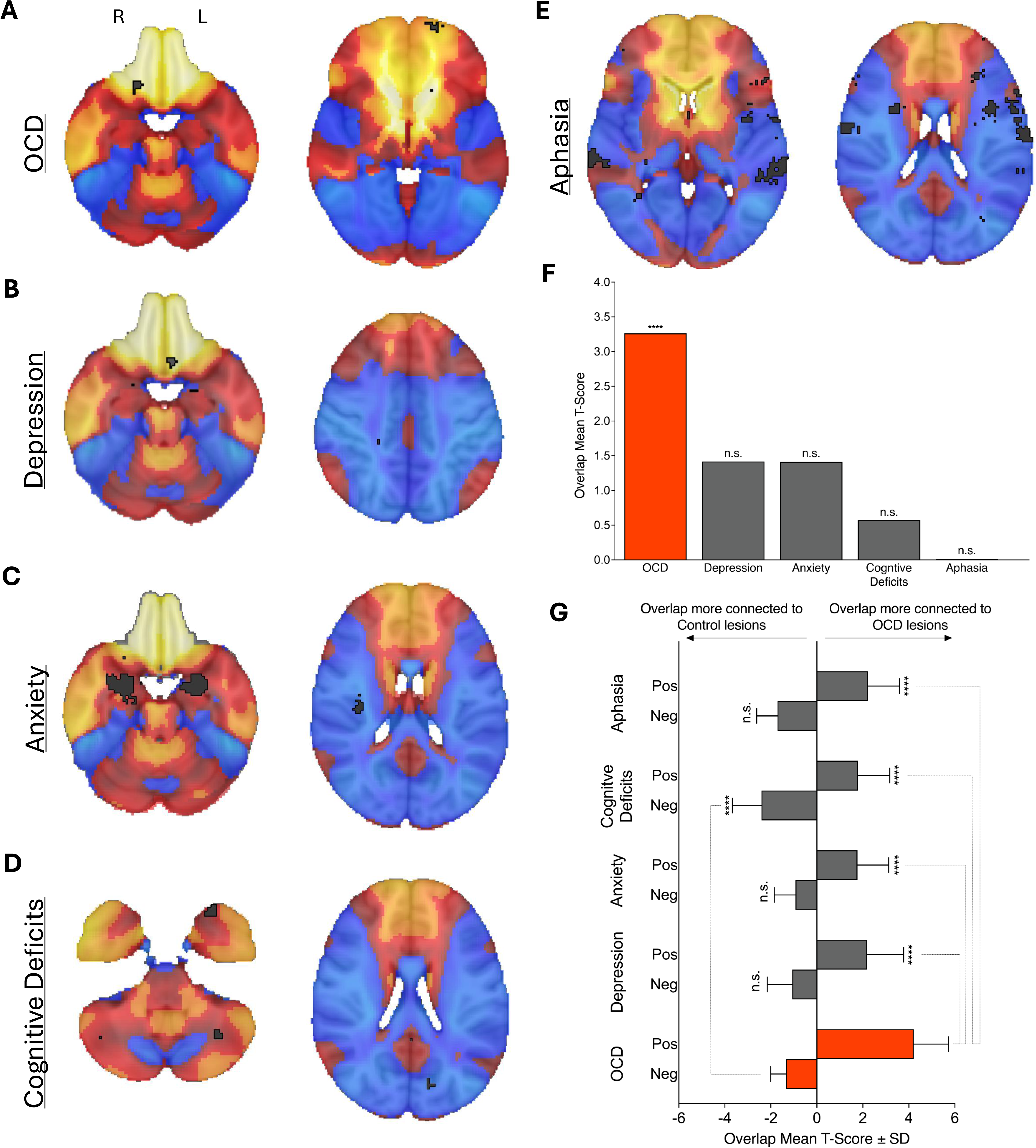
Lesional OCD connectivity overlaps with functional connectivity nodes associated with OCD. We tested the association of lesional OCD connectivity to OCD functional peaks associated with the term “OCD”, obtained from the metanalytic tool Neurosynth^36^. In the map slices, warm and cold colors represent areas that are more or less connected to OCD lesions when compared to control lesions, respectively, in the Lesional OCD vs. Controls Connectivity Map, while functional peaks are overlapped in black. The functional peaks associated to “OCD” (**A**) “depression” (**B**), “anxiety” (**C**), “cognitive deficits” (**D**) and “aphasia” (**E**) are represented. **F.** In an ROI wise approach, OCD functional ROIs overlapped more strongly with the Lesional OCD vs. Controls Connectivity Map than with randomly permuted maps (p<0.001). This was not found for any of the control ROIs p>0.05 for all, Bonferroni-corrected permutation tests;). **G.** In a voxel-wise approach, we confirmed that the overlap with “OCD” functional ROIs occurs particularly in voxels that are more connected to OCD lesions (positive T values of the Lesional OCD vs. Controls Connectivity Map), that had significantly higher mean T-values than voxels in functional ROIs for the other tested symptoms (p<0.001 for all, Bonferroni-corrected t-tests). For voxels that are more connected to control lesions (negative T values of the Lesional OCD vs. Controls Connectivity Map), differences relative to “OCD” functional ROIs were not statistically significant relative to those of other symptoms (p>0.05), with the exception of “Cognitive Deficits” (p<0.001, Bonferroni-corrected t-tests). **** – p-value<0.001; n.s. – not significant; OCD – obsessive compulsive disorder.

In addition to the meta-analytic approach using Neurosynth, we also used resting-state fMRI data from a sex and age-matched case-control study of primary OCD ongoing in our group. From an initial pool of 201 subjects, 54 patients with OCD and 61 healthy participants, with non-significant differences in age (p=0.60) and sex (p=0.97), were eligible and had fMRI scans available for this analysis (Figure 5A). Using a hypothesis-driven approach, we used MDMR analysis^37–40,59^ to test if the pattern of whole brain functional connectivity to a seed region was different between patients with OCD and healthy volunteers. The seed was the intersection of the Lesional OCD Mean Connectivity Map and the Lesional OCD vs. Controls Connectivity Map (Figure 3C), as these regions reflect those with the greatest and most specific connectivity to lesions causing OCD. We found that, in primary OCD, the connectivity to lesional OCD intersection regions was different from that of healthy subjects (p=0.01; Figure 5B), even when controlling for age, sex, and antidepressant equivalent dose (p=0.02). Finally, in an exploratory analysis, we identified that connectivity to the regions of the lesional OCD intersection map (Figure 3C; Figure 5B, light green areas) differed significantly between OCD and healthy participants across several ROIs (Figure 5C). We found significant differences in the left caudate (T=4.5), left frontal pole (T=-5.5; Figure 5C), left precentral gyrus (T=5.2; Figure 5C), left supramarginal gyrus (T=-4.5; Figure 5C), right frontal gyrus (T=-6.4 and T=-4.9) and right frontal pole (T=-4.99; p<0.0001 for all, Bonferroni corrected two-sample t-tests).

**Figure 5.**
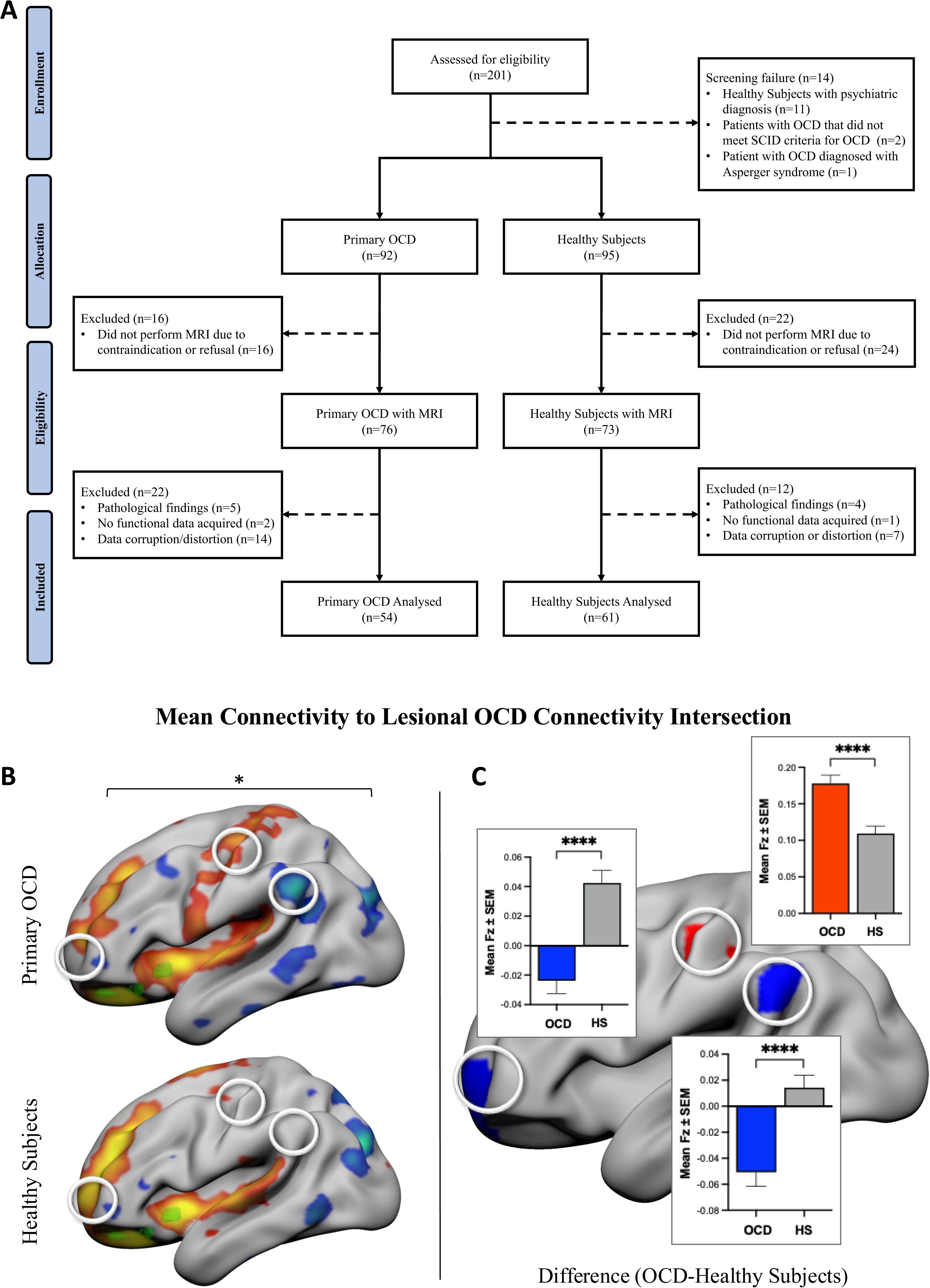
Connectivity to the lesional OCD circuit is disrupted in primary OCD. **A.** From an ongoing age and sex frequency-matched case-control study conducted at the Champalimaud Foundation, resting state fMRI data from 54 patients with primary OCD and 61 healthy volunteers was available for analysis. **B.** Whole-brain connectivity to a seed region reflecting the Lesional OCD Intersection Map (Figure 3C) differed between patients with primary OCD and healthy controls. Here we represent mean surface connectivity to the intersection region (Fz) of patients with OCD (top panel) and healthy volunteers (lower panel), to illustrate the differences. In both maps, warm and cold colors represent regions respectively more positively or negatively connected to the lesional OCD connectivity intersection map, that is represented in green. **C.** In an exploratory approach, we identified ROIs >120 mm³ with contiguous voxels with a between-group difference >0.06 Fz for mean connectivity to the lesional OCD connectivity intersection map, in the same direction. Here, left hemisphere lateral view is presented to show three examples of such brain regions, namely the precentral gyrus, significantly more connected to the intersection region in patients diagnosed with primary OCD, and the supramarginal gyrus and frontal pole, significantly more anti-correlated to the intersection region in patients diagnosed with primary OCD when compared to healthy subjects (Bonferroni-corrected two-sample t-tests). * – p-value<0.05; **** – p-value<0.001; HS – Healthy Subjects; OCD – obsessive compulsive disorder; ROI – region of interest.

Lastly, we explored whether the functional connectivity profiles emerging from our lesional OCD analysis intersected with well-known effective targets used for DBS and TMS neuromodulatory treatments for primary OCD. We found that the four different DBS targets reported to be effective for primary OCD symptoms (ALIC, amSTN, Nac, and VC-VS) all fall within lesion-based circuits, in both the sensitivity map for lesional OCD connectivity (Figure 6A) and the specificity map for lesional OCD connectivity (Figure 6B). Similarly, the brain region that is targeted by TMS devices cleared and/or approved by international regulatory agencies for treatment of OCD (mPFC-ACC) also intersected with lesion-based circuits (Figure 6A & B). Indeed, the mPFC-ACC region has been targeted using two distinct TMS coils, for which different electric fields have been computed^60^. Interestingly, while the two electric fields have very different sizes, both overlap with the most positive regions of the two connectivity maps in dorsal regions of the frontal cortex (Figure 6C-F).

**Figure 6.**
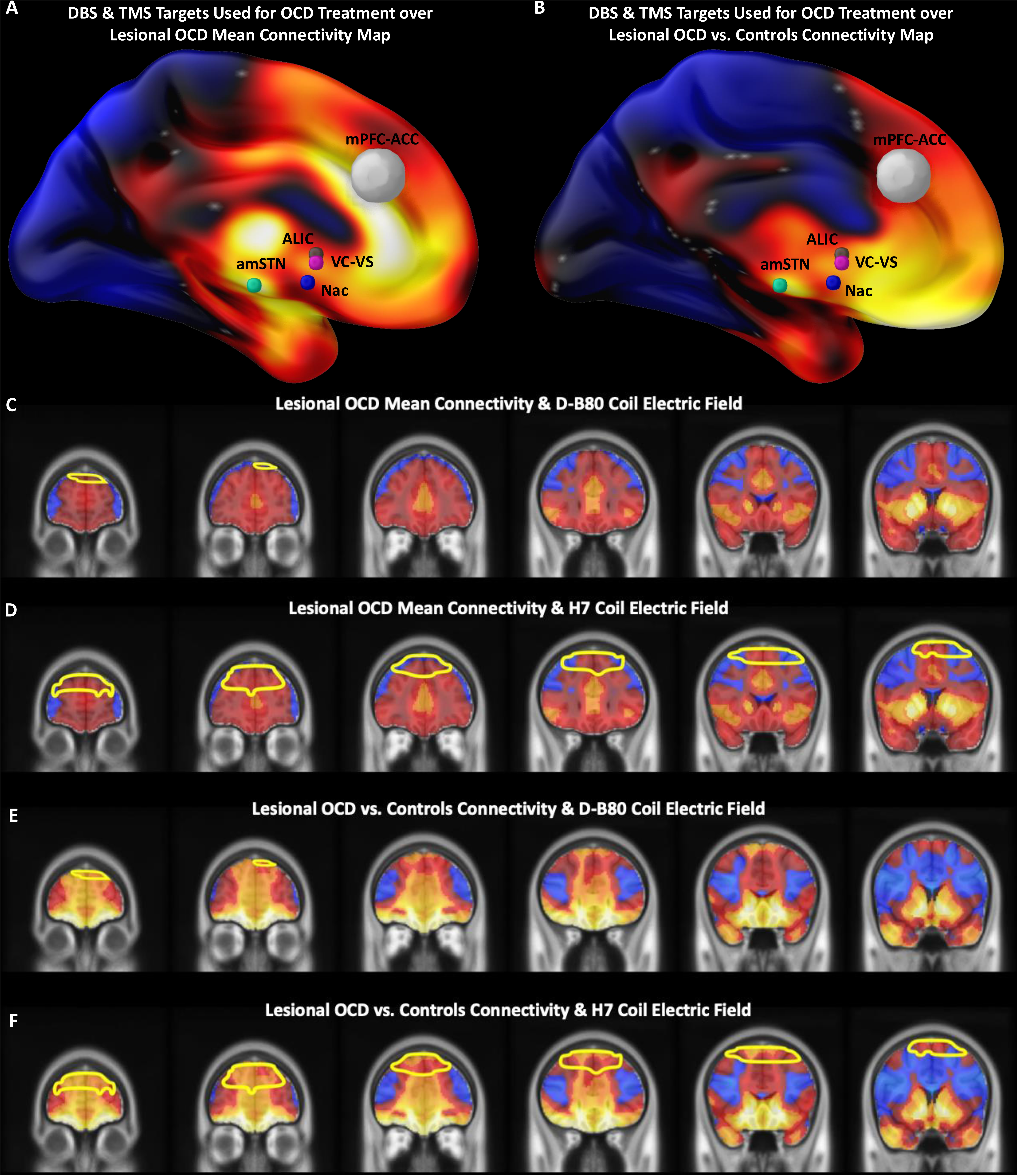
Therapeutic implications of lesional OCD functional connectivity. **A.** DBS targets used in OCD treatment, namely ALIC (grey), amSTN (green), Nac (blue) and VC-VS (violet), and the mPFC-ACC target used for TMS (white) are aligned with regions positively connected to OCD lesions, within the Lesional OCD Mean Connectivity Map. **B.** Similarly, these targets aligned with regions more specifically connected to OCD lesions in the Lesional OCD vs. Controls Connectivity Map. Two TMS coils have been used to target the mPFC-ACC target, namely the D-B80 coil and the H7 coil, with very distinct electric fields^60^. Here we show overlap of the Lesional OCD Mean Connectivity Map with the electric fields (yellow outlines, as adapted from a previous report^60^) of the D-B80 coil (**C**) and the H7 coil (**D**), as well as overlap of the Lesional OCD vs. Controls Connectivity Map with the electric fields of the D-B80 coil (**E**) and the H7 coil (**F**). In the sensitivity maps represented in A, C and D, warm and cold colors represent areas that are positively or negatively connected to OCD lesions, respectively. On the other hand, in the specificity maps represented in B, E and F, the warm and cold colors represent areas that are more or less connected to OCD lesions when compared to controls, respectively. ACC – anterior cingulate cortex; ALIC – anterior limb of internal capsule; amSTN – anteromedial subthalamic nucleus; mPFC – medial prefrontal cortex; Nac – nucleus accumbens; OCD – obsessive compulsive disorder; – VC-VS – ventral capsule/ventral striatum.

## DISCUSSION

While in most cases of OCD no specific etiology can be identified, this neuropsychiatric disorder may develop after brain insults which, as expected and confirmed here, has a late onset (mean age 46.4±16.9 years) when compared with primary OCD (19.5±45.5 years, according to Ruscio et al.^56^), and even late onset primary OCD (46.4±16.9 vs. 24.9±9.3 years, according to Anholt et al.^61^). We show that while occurring in a variety of different locations, brain lesions associated to OCD are more frequently located in the temporal lobes, basal ganglia and frontal lobes, with the OFC, bilaterally, and the right temporal pole more frequently affected that in control lesions. Importantly, the normative functional connectivity of lesion locations associated with OCD was quite distributed, including peak connectivity with the basal ganglia, bilaterally, with a unique pattern of brain connectivity to the OFC and ventral basal ganglia, also bilaterally, in comparisons with control lesions. This OCD lesion network map (LNM) was found to be robust to several potential confounders, including lesion etiology, beyond what was found for lesional topography maps. Moreover, the OCD LNM overlapped with known brain functional connectivity properties of primary OCD, revealed abnormal connectivity patterns in an independent primary OCD cohort, and aligned well with therapeutic brain stimulation targets for primary OCD.

Similarly to what we have described here for OCD, over the last century neuropsychiatric disorders secondary to brain insults have been open windows to probe the neurobiology, particularly the neuroanatomy, of complex neurologic and psychiatric conditions^62^. They allow for clearer inferences regarding causal associations between brain regions and behavioral changes, potentially revealing areas and networks that may be overlooked when studying idiopathic or primary neuropsychiatric conditions^63^. To the best of our knowledge, while different authors have described the occurrence of OCD symptoms due to a brain insult^3–5^, this is the first time lesional OCD was studied using a systematic approach, disentangling its neuroanatomic substrate. In this study, we found that OCD symptoms are associated to cortico-striato-thalamo-cortical (CSTC) circuit dysfunction^64–67^, particularly in the OFC and ventral basal ganglia, bilaterally. Both regions, when lesioned, have been associated to deficits in cognitive functions^67^ that have also been described in patients with primary idiopathic OCD, such as response inhibition, set shifting and decision-making^67,68^.

OCD-related lesions were preferentially located in frontal regions as well as the basal ganglia. However, OCD lesion topography lacked robust specificity and is significantly influenced by the nature of the lesions, namely vascular vs. tumoral lesions. In contrast, OCD lesional connectivity was specific for the OFC and ventral basal ganglia bilaterally, and proved to be more robust to controls for lesion etiology. Regions of specific OCD lesional connectivity were further refined by thresholding of the Lesional OCD vs. Controls Connectivity Map and intersection with the Lesional OCD Mean Connectivity Map, revealing peak connectivity regions in the frontal pole, frontal medial and orbital cortices and ventral basal ganglia, bilaterally. Importantly, auto-connectivity from lesion locations alone was not sufficient to explain the lesional OCD peak connectivity pattern, since exclusion of lesions that overlapped with brain regions of peak OCD lesional connectivity did not change the lesional connectivity pattern. This lesional OCD network has some commonality with other lesional networks. The functional networks of lesional mania also included the OFC, albeit only the right hemisphere^9^, and this frontal region was also an important hub in lesions associated to criminal behaviour^29^. This common neuroanatomic substrate is not surprising, since the OFC has been associated to cognitive functions, such as decision making and reward appraisal, that are commonly impaired in several neuropsychiatric conditions, including OCD^69–72^. On the other hand, ventral basal ganglia may be more specific for OCD symptoms. In fact, lesions of the basal ganglia have been more frequently associated to movement disorders, namely parkinsonism^73^, dystonia^74^ and tic disorder^75^. However, even in tic disorder^75^, a condition highly co-morbid with OCD^76^, this was mainly restricted to dorsal regions of the basal ganglia. Indeed, the ventral striatum is one of several potential targets for DBS treatment of OCD^77^, and structural connectomics of OCD-DBS targets have suggested that therapeutic effects reflect circuit neuromodulatory effects in ventral fronto-striato-thalamic pathways involving the ventromedial prefrontal cortex, OFC and ventral basal ganglia^78,79^. Future studies comparing the neuroanatomic substrate of lesional OCD with other lesional neuropsychiatric syndromes may provide further insights into the specific functional neuroanatomy of OCD, and contribute to development of new or optimized treatments.

Regions highlighted not only by the topographic, but mainly by the connectivity maps of lesional OCD are aligned with many of the findings of the numerous case-control neuroimaging studies that have been conducted in primary OCD^65–67^. Meta-analyses of those studies have found cortical and subcortical volume abnormalities in patients with OCD, mainly involving the OFC and basal ganglia, but also the hippocampus, anterior cingulate cortex, and thalamus^65,66^. The OFC in particular has been consistently reported to have decreased global and GM volume^67^, while for the basal ganglia the direction of findings has been less consistent, possibly reflecting differences in the definition of regions of interest or heterogeneity in OCD phenotype^67^. Nevertheless, structural evidence in the basal ganglia is stronger in ventral structures, namely ventral putamen or nucleus accumbens^67,80,81^, which is consistent with the ventral striatum being a target for DBS in OCD^80^. Again, this is aligned with our findings, since the ventral caudate, ventral putamen and nucleus accumbens were found to be highly significant nodes of the lesional OCD network, bilaterally. Additionally, spectroscopy studies^82^ and other functional neuroimaging studies, such as symptom provocation during fMRI or positron emission tomography^67^, have also shown altered metabolism and dysfunctional activity in CSTC loops, including in OFC and basal ganglia^67,82^. The findings described in cross-sectional and case-control studies of primary OCD should be regarded as primarily correlational and are certainly confounded by factors such as medication and comorbidity^65–67^.Our results add empirical weight to the hypothesis that structural and functional abnormalities observed in these regions in the context of primary OCD may be causally related to occurrence of the disorder.

Indeed, here we specifically explored the potential relationships between lesional and primary OCD, i.e., in patients with an OCD diagnosis that was not caused by a known brain lesion or any other specific etiology. First, we confirmed that functional regions associated with primary OCD significantly overlapped with the lesional OCD connectivity profile, an effect that was not found for any of the other neuropsychiatric syndromes that we studied using the same method. Second, we also tested whether lesional OCD functional connectivity parallels abnormal connectivity in primary OCD. In data collected within our own fMRI study of primary OCD with a case-control design, we found that, in patients with primary OCD, the whole-brain connectivity profile to lesional OCD connectivity nodes was different from the profile found in healthy subjects. In subsequent exploratory analyses, we found that in patients diagnosed with primary OCD, when compared to healthy volunteers, the lesional OCD nodes were differently connected to the left precentral and supramarginal gyrus, left putamen, right frontal middle gyrus and frontal pole, bilaterally. These findings further support that the dysconnectivity profile that we described in post-lesional OCD may in fact be a common substrate for OCD, even in the absence of a brain lesion. While promising, the relationship between lesional and primary OCD neuroimaging findings should be explored in future studies, to further contribute towards a better understanding of OCD pathophysiology.

Interestingly, our lesional OCD connectivity profile may also have potential implications in OCD treatment, when considering therapeutic strategies for focal neuromodulation. Indeed, we found that different targets across different stimulation modalities overlap positive connectivity nodes of the lesional OCD connectivity profile in both the sensitivity connectivity map (Lesional OCD Mean Connectivity) and the specificity map (Lesional OCD vs. Controls Connectivity). In fact, our results may help reconcile the somewhat inconsistent evidence concerning the two TMS coils cleared by the Food and Drug Administration for treatment of OCD. While both coils have been shown to be effective^47,83^, their electric fields are distinct, namely in size^60^. Interestingly, despite such a difference, the two electric fields overlapped some of the most positive regions of the sensitivity and specificity lesional connectivity maps, suggesting that both coils are targeting clinically relevant dysfunctional OCD networks. This is not the first time that lesional connectivity maps obtained from lesional neuropsychiatric disorders have been useful in guiding brain stimulation strategies in neuropsychiatric syndromes^9,73,84–87^. Lesional OCD connectivity profiles may thus contribute to refine current targets or identify additional brain stimulation targets to be tested in the future. Nevertheless, and while promising, the impact of our results in guiding neuromodulation strategies should be viewed with caution, as clinical validation is warranted.

These results should be interpreted considering potential limitations. First, the lesional OCD cohort was extracted from literature cases. Consequently, it may be susceptible to various biases. Indeed, analyses were performed using a limited number of slices from each scan, that did not encompass the whole lesion, and lesions were represented using heterogenous imaging modalities, increasing the likelihood of errors in lesion-tracing. However, lesion maps obtained from limited numbers of slices per lesion have already shown to yield consistent results when compared to those obtained from whole lesion cohorts^8,9,25^. Moreover, when formally testing the robustness and reliability of the results obtained from the topography and functional connectivity analyses, we have shown that neither the authors’ neuroimaging representation modalities nor the lesion tracing methods impacted our results. Another potential limitation, inherent to all lesion studies, concerns the fact that such studies ignore the impact of time in the post-lesional brain, particularly the impact of tissue remodeling that occurs in response to any brain insult. Ultimately, these neurobiological mechanisms may lead to activity changes in regions of the brain that are not connected to the lesion location and that are not accounted for when performing topography and lesion network mapping analyses. Such limitation is difficult to overcome using the current study design, and future prospective research should be conducted to help clarify the role of post-lesional changes in brain architecture and how they impact the functional connectivity profile of the post-lesional OCD brain. Nevertheless, it is reassuring that we found that lesional OCD “disconnections” were paralleled by a converging dysfunctional connectivity profile in primary OCD, suggesting a common substrate for OCD symptoms, irrespective of their etiology. An additional limitation, common to lesion network mapping studies^88,89^, concerns thresholding of connectivity maps to obtain the intersection map. While the threshold definition was observer dependent, to mitigate this limitation, all thresholds were defined following a specific rationale (please see *Methods*) and resulted from a consensual decision by a group of researchers. Lastly, we cannot exclude that, in some of the cases included in our control brain lesion sample, OCD symptoms could be present. Nevertheless, we expect the impact of such cases to be negligible. Lesional OCD is uncommon, and our control sample is large (N=608) and heterogeneous (vascular and tumor lesions), to ensure adequate comparisons while achieving appropriate statistical power, as has been recommended^90^. Moreover, the presence of false negative patients in the control sample would, if anything, bias the result towards the null hypothesis.

In conclusion, while brain lesions causing OCD-syndromes may occur in a variety of different brain locations, they are not randomly distributed in the brain. Specifically, OCD occurring after a brain lesion is specifically associated to locations involving the OFC bilaterally, as well as the right temporal pole, and converge on specific brain networks connected to the OFC and ventral basal ganglia, bilaterally. These results emphasize the critical role of these structures in the neurobiology of OCD, providing additional empirical weight to the hypothesis that structural and functional abnormalities observed in these structures in the context of primary OCD are causally related to the disorder and its symptoms. Moreover, the lesional OCD connectivity network described here may prove helpful in guiding brain stimulation strategies with potential therapeutic effects for this neuropsychiatric disorder.

## AUTHORS CONTRIBUTIONS

GC, JBB-C, and AJO-M conceived and designed the work; GC, ND, CF, JR, AM, JO, NL, JBB-C, and AJO-M acquired the data; GC, ND, JC-I, DM, FFV, CF, NL, SHS, MDF, JBB-C, and AJO-M analyzed and interpreted data; GC and AJO-M drafted the work; ND, JC-I, DM, FFV, CF, JR, AM, JO, NL, SHS, MDF, and JBB-C revised the manuscript critically for important intellectual content. All authors approved the final version to be published and agree to be accountable for all aspects of the work in ensuring that questions related to the accuracy or integrity of any part of the work are appropriately investigated and resolved. GC and AJO-M had full access to all the data in the study and take responsibility for the integrity of the data and the accuracy of the data analysis.

## DATA AVAILABILITY STATEMENT

The data supporting the conclusions of this article are not readily available but may be accessed upon reasonable request to the corresponding author.

## ETHICS STATEMENT

The study was conducted in accordance with the Declaration of Helsinki and was approved by the Champalimaud Foundation Ethics Committee. Written informed consent was obtained from all recruited participants.

## ACKNOWLEDGMENTS

We would like to acknowledge the contribution of Julia de Souza Queiroz to data collection. The normative connectome was provided by the Human Connectome Project, WU-Minn Consortium (Principal Investigators: David Van Essen and Kamil Ugurbil; 1U54MH091657) funded by the 16 NIH Institutes and Centers that support the NIH Blueprint for Neuroscience Research; and by the McDonnell Center for Systems Neuroscience at Washington University.

