## Supplemental for "Obsessive-compulsive disorder secondary to focal brain lesions: from lesions to networks"

**SUPPLEMENTARY METHODS**

**A - Supplementary methods for lesional OCD cohort and analyses**

1) **Inclusion and exclusion criteria for lesional OCD cohort**

Inclusion criteria:

- Documented history of obsessive-compulsive symptoms or related disorders. Obsessions are defined as recurrent and persistent thoughts, urges or images that are intrusive, unwanted and cause marked anxiety, while compulsions are repetitive behaviors or mental acts that the person feels driven to perform in response to an obsession, or according to the rules that must be applied rigidly. Reports that do not provide details on behavioral changes remain eligible if authors stated that they met contemporary diagnostic criteria;
- Any brain lesion regardless of its nature, location, size and age of occurrence, as long as occurring before the first manifestation of obsessive-compulsive (or related disorders) symptoms and/or signs. Cases where injury has been diagnosed after the first manifestations may still be considered eligible if there is unequivocal evidence that the lesion was acquired before the psychiatric manifestations developed.
- Brain lesion images collected after the age of 18 years-old;

Exclusion criteria:

- Patients in whom a presumed brain lesion has not been confirmed by in vivo imaging or post-mortem methods;
- Patients in whom the obsessive-compulsive episode may have been induced by factors or events other than a structural brain-insult.

2) **Additional preprocessing steps performed in the Human Connectome Project neuroimage data**

Lesion network maps were generated using resting state functional magnetic resonance imaging (rs-fMRI) data from the Human Connectome Project, an openly available database including high resolution 3T and 7T MRI data as well as MEG and EEG data^1^. As described in Glasser et al.^2^, minimally preprocessed 3T rs-fMRI data from 937 healthy young adults were extracted from the database to perform the analysis. This dataset was further processed in accordance with Fox et al. 2005^3^. Scans were temporally filtered below and above 0.08Hz and 0.009Hz, respectively, and spatially smoothed using a Gaussian kernel of 6 mm full-width half-maximum. Then, nonspecific sources of variability were removed through regression of six movement parameters resulting from the preprocessing pipeline, mean signal from whole brain, ventricles, and deep white matter masks, as well as their first order derivatives. Additionally, to guaranty that steady-state magnetization was achieved, the first 10 volumes were removed from the data^4^.

**A - Supplementary methods for primary OCD case-control study and data analyses**

1) **Inclusion and exclusion criteria for the primary OCD and healthy volunteer groups**

Inclusion criteria:

- Age between 18 and 65 years-old;
- Ability to provide informed consent;
- Fluent in Portuguese or English;
- Established diagnosis of OCD according to Structured Clinical Interview for the Diagnostic and Statistical Manual of Mental Disorders 5^th^ Edition (SCID-5) – for the primary OCD group only;
- Does not meet DSM criteria for any of the psychiatric diagnoses screened by the Mini International Neuropsychiatric Interview – for the healthy subjects group only.

Exclusion criteria:

- Any acute medical illness;
- Substance abuse or dependence in the last 12 months;
- Pregnancy;
- Dementia, developmental disorders with low intelligence quotient or any other form of cognitive impairment;
- Active neurological disease;
- Previously known or newly identified structural lesion of the central nervous system;
- Illiteracy or otherwise not understanding the study’s instructions;
- Psychotic or mood disorder condition requiring hospitalization at the moment of recruitment.

2) **rs-fMRI acquisition and preprocessing steps**

Sequences were acquired on a 3T scanner (Ingenia, Philips Healthcare, Best, The Netherlands) with 32-channel head coil. Volumetric T1-weighted Fast Field Echo (3DT1-FFE) (time of echo (TE)/time of repetition (TR)= 2.7/6.2ms; inversion time (TI)=950ms; FOV=240x240x180 mm; matrix=240x240; isometric voxel=1mm^3^). Functional connectivity during rest (with eyes open and a fixation cross) was acquired with voxel size: 3x3x3 mm, TE/TR: 30/2280, FOV: 240x240, slice thickness: 3mm, 200 dynamic scans. Functional data were preprocessed with the CONN toolbox (<http://www.nitrc.org/projects/conn>; pipeline: direct registration with fieldmap)^5^, including motion estimation, fieldmap-based susceptibility distortion correction, slice timing correction, transformation to Montreal Neurological Institute (MNI) space template (voxel size of 2x2x2 mm). This pipeline labels time points with framewise displacement greater than 0.9 mm or global signal changes above 5 standard deviations as potential outliers. Preprocessing also included the extraction of tissue probability maps (TPMs) for gray matter (GM), white matter (WM) and cerebrospinal fluid (CSF) from the subject’s T1w image. The preprocessed functional images were further denoised with in-house developed tools, including temporal high-pass filtering, spatial smoothing, nuisance regression and scrubbing. First, to remove slow trends, data was filtered with an ideal high-pass filter with 0.01Hz cutoff frequency. Then spatial smoothing was applied using a Gaussian kernel with a full-width-at-half-maximum of 4 mm, followed by nuisance regression, including the 6 subject motion parameters, their 6 derivatives, 12 principal components (PCs) from WM and CSF, and non-valid time points (3 first non-steady-state volumes and motion outliers). PCs were extracted by principal component analysis of the BOLD time series from within a mask of the union of CSF and WM generated from the TPM segmentation, from which all subcortical structures (as from Harvard-Oxford atlas) were masked out and which was subsequently eroded to minimize partial-volume contamination from GM. PC extraction was performed after filtering and before smoothing, running smoothing within the eroded WM+CSF mask to avoid further contamination from GM. The motion regressors were also high-pass filtered to avoid re-introduction of low frequency trends^6,7^. We included time points to be scrubbed in the design matrix to avoid non-valid volumes to affect the estimation, namely the first RF-excited time points (5 volumes/13.5 s were excluded), and outliers detected in the preprocessing stage. We conducted a thorough quality control on the acquired, preprocessed and denoised functional images on a subject-by-subject basis. Functional runs with imaging artifacts or issues not removed in the denoising step, as well as those with insufficient valid time points (less than 112 volumes/5 min), were discarded for further analysis.

**SUPPLEMENTARY TABLES**

**Table S1 –** Search syntax

|  | **Psychiatric Disorder** | **Brain** | **Insult** | **Not Topic** |
| --- | --- | --- | --- | --- |
| **Pubmed*** | Obsessive-Compulsive | Cerebral | Lesion |  |
|  | Obsessive Compulsive | Cerebellum | Injury |  |
|  | OCD | Brain | Tumor |  |
|  | Obsessive | Central Nervous System | Neoplasm |  |
|  | Compulsive | CNS | Mass |  |
|  | Obsession |  | Infection |  |
|  | Compulsion |  | Abscess |  |
|  | Rumination |  | Cyst |  |
|  | Hoarding |  | Stroke |  |
|  | Trichotillomania |  | Hemorrhage |  |
|  | Excoriation |  | Bleeding |  |
|  | Body Dysmorphic |  |  |  |
|  | Anxiety Disorder |  |  |  |
| **Web of Science**** | Obsessive-Compulsive | Cerebral | Lesion | Animal |
|  | Obsessive Compulsive | Cerebellum | Injury | Monkey |
|  | OCD | Brain | Tumor | Chimpanzee |
|  | Obsessive | Central Nervous System | Neoplasm | Mouse |
|  | Compulsive | CNS | Mass | Mice |
|  | Obsession |  | Infection | Rat |
|  | Compulsion |  | Abscess | Cat |
|  | Rumination |  | Cyst | Dog |
|  | Hoarding |  | Stroke | Rabbit |
|  | Trichotillomania |  | Hemorrhage | Bird |
|  | Excoriation |  | Bleeding | Fish |
|  | Body Dysmorphic |  |  | Child |
|  | Anxiety Disorder |  |  |  |
| **EMBASE***** | Obsessive-Compulsive | Cerebral | Lesion |  |
|  | Obsessive Compulsive | Cerebellum | Injury |  |
|  | OCD | Brain | Tumor |  |
|  | Obsessive | Central Nervous System | Neoplasm |  |
|  | Compulsive | CNS | Mass |  |
|  | Obsession |  | Infection |  |
|  | Compulsion |  | Abscess |  |
|  | Rumination |  | Cyst |  |
|  | Hoarding |  | Stroke |  |
|  | Trichotillomania |  | Hemorrhage |  |
|  | Excoriation |  | Bleeding |  |
|  | Body Dysmorphic |  |  |  |
|  | Anxiety Disorder |  |  |  |
| **PsycINFO ****** | Obsessive-Compulsive | Cerebral | Lesion |  |
|  | Obsessive Compulsive | Cerebellum | Injury |  |
|  | OCD | Brain | Tumor |  |
|  | Obsessive | Central Nervous System | Neoplasm |  |
|  | Compulsive | CNS | Mass |  |
|  | Obsession |  | Infection |  |
|  | Compulsion |  | Abscess |  |
|  | Rumination |  | Cyst |  |
|  | Hoarding |  | Stroke |  |
|  | Trichotillomania |  | Hemorrhage |  |
|  | Excoriation |  | Bleeding |  |
|  | Body Dysmorphic |  |  |  |
|  | Anxiety Disorder |  |  |  |

* We applied the following filters: Humans; age +19; English, Portuguese, Spanish, German and French.

** We applied the following filters: English, Portuguese, Spanish, German and French.

*** We applied the following filters: Humans; Young adults, adult, middle aged, aged and very elderly.

**** We applied the following filters: Adults; 18 or older; English, Portuguese, Spanish, German and French.

**Table S2 –** Comparison of Right vs. Left Hemisphere Lesions in lesional OCD-like symptoms cases

| **Brain Areas** | **% of lesioned voxels (N=40)** | | | | | | |
| --- | --- | --- | --- | --- | --- | --- | --- |
|  | **Left** | | | **Right** | | | **P Value (Sign Test)^a^** |
|  | **Min** | **Median** | **Max** | **Min** | **Median** | **Max** |  |
| **Automated Anatomical Labeling Atlas** | | | | | | | |
| **Frontal** | | | | | | | |
| precentral | 0 | 0 | 1.32 | 0 | 0 | 1.31 | 0.63 |
| frontal_sup_2 | 0 | 0 | 8.56 | 0 | 0 | 5.05 | 1.00 |
| frontal_mid_2 | 0 | 0 | 2.12 | 0 | 0 | 0.82 | 0.34 |
| frontal_inf_oper | 0 | 0 | 1.64 | 0 | 0 | 0.79 | 0.38 |
| frontal_inf_tri | 0 | 0 | 4.68 | 0 | 0 | 1.14 | 1.00 |
| frontal_inf_orb_2 | 0 | 0 | 15.38 | 0 | 0 | 14.15 | 0.69 |
| rolandic_oper | 0 | 0 | 19.86 | 0 | 0 | 5.97 | 1.00 |
| supp_motor_area | 0 | 0 | 22.31 | 0 | 0 | 9.31 | 1.00 |
| olfactory | 0 | 0 | 12.50 | 0 | 0 | 10.38 | 0.45 |
| frontal_sup_medial | 0 | 0 | 9.69 | 0 | 0 | 12.02 | 1.00 |
| frontal_med_orb | 0 | 0 | 19.12 | 0 | 0 | 17.89 | 1.00 |
| rectus | 0 | 0 | 21.29 | 0 | 0 | 20.81 | 0.29 |
| ofcmed | 0 | 0 | 21.69 | 0 | 0 | 20.78 | 1.00 |
| ofcant | 0 | 0 | 5.62 | 0 | 0 | 12.00 | 0.22 |
| ofcpost | 0 | 0 | 12.71 | 0 | 0 | 16.41 | 1.00 |
| ofclat | 0 | 0 | 1.02 | 0 | 0 | 7.52 | 1.00 |
| **Cingulum** | | | | | | | |
| cingulate_ant | 0 | 0 | 13.64 | 0 | 0 | 7.84 | 1.00 |
| cingulate_mid | 0 | 0 | 12.78 | 0 | 0 | 4.36 | 0.38 |
| cingulate_post | 0 | 0 | 3.46 | 0 | 0 | 1.19 | 1.00 |
| **Subcortical Grey Matter** | | | | | | | |
| caudate | 0 | 0 | 14.83 | 0 | 0 | 7.25 | 1.00 |
| putamen | 0 | 0 | 25.07 | 0 | 0 | 8.08 | 0.45 |
| pallidum | 0 | 0 | 24.57 | 0 | 0 | 13.93 | 0.75 |
| thalamus | 0 | 0 | 9.82 | 0 | 0 | 3.22 | 0.45 |
| **Occipital** | | | | | | | |
| calcarine | 0 | 0 | 1.76 | 0 | 0 | 4.89 | 0.25 |
| cuneus | 0 | 0 | 0.27 | 0 | 0 | 1.26 | 1.00 |
| lingual | 0 | 0 | 2.39 | 0 | 0 | 7.55 | 1.00 |
| occipital_sup | 0 | 0 | 0 | 0 | 0 | 1.25 | 1.00 |
| occipital_mid | 0 | 0 | 0.31 | 0 | 0 | 8.34 | 1.00 |
| occipital_inf | 0 | 0 | 0 | 0 | 0 | 11.36 | 1.00 |
| fusiform | 0 | 0 | 0.30 | 0 | 0 | 11.87 | 0.22 |
| **Parietal** | | | | | | | |
| postcentral | 0 | 0 | 4.90 | 0 | 0 | 0.09 | 1.00 |
| parietal_sup | 0 | 0 | 4.38 | 0 | 0 | 0 | 1.00 |
| parietal_inf | 0 | 0 | 1.02 | 0 | 0 | 0 | 0.50 |
| supramarginal | 0 | 0 | 1.83 | 0 | 0 | 1.66 | 1.00 |
| angular | 0 | 0 | 1.88 | 0 | 0 | 0 | 0.50 |
| precuneus | 0 | 0 | 5.25 | 0 | 0 | 3.60 | 1.00 |
| paracentral_lobule | 0 | 0 | 9.08 | 0 | 0 | 0 | 1.00 |
| **Insula and Temporal** | | | | | | | |
| insula | 0 | 0 | 9.80 | 0 | 0 | 12.03 | 1.00 |
| hippocampus | 0 | 0 | 2.58 | 0 | 0 | 21.88 | 0.07 |
| parahippocampal | 0 | 0 | 1.99 | 0 | 0 | 16.87 | 0.69 |
| amygdala | 0 | 0 | 0 | 0 | 0 | 31.45 | 0.25 |
| heschl | 0 | 0 | 2.22 | 0 | 0 | 6.15 | 1.00 |
| temporal_sup | 0 | 0 | 13.35 | 0 | 0 | 13.61 | 0.13 |
| temporal_pole_sup | 0 | 0 | 5.53 | 0 | 0 | 16.74 | 0.75 |
| temporal_mid | 0 | 0 | 9.07 | 0 | 0 | 15.25 | 0.11 |
| temporal_pole_mid | 0 | 0 | 0 | 0 | 0 | 13.08 | 0.06 |
| temporal_inf | 0 | 0 | 3.23 | 0 | 0 | 8.43 | 0.07 |
| **Cerebellum** | | | | | | | |
| cerebelum_crus1 | 0 | 0 | 0.79 | 0 | 0 | 0 | 1.00 |
| cerebelum_crus2 | 0 | 0 | 3.80 | 0 | 0 | 0 | 1.00 |
| cerebelum_3 | 0 | 0 | 0 | 0 | 0 | 1.46 | 1.00 |
| cerebelum_4_5 | 0 | 0 | 0.94 | 0 | 0 | 2.87 | 1.00 |
| cerebelum_6 | 0 | 0 | 1.92 | 0 | 0 | 0.18 | 1.00 |
| cerebelum_7b | 0 | 0 | 7.25 | 0 | 0 | 0 | 1.00 |
| cerebelum_8 | 0 | 0 | 1.37 | 0 | 0 | 0 | 1.00 |
| cerebelum_9 | 0 | 0 | 0 | 0 | 0 | 0 | 1.00 |
| cerebelum_10 | 0 | 0 | 0 | 0 | 0 | 0 | 1.00 |
| **John Hopkins University Atlas** | | | | | | | |
| **White Matter** | | | | | | | |
| corticospinal_tract | 0 | 0 | 0 | 0 | 0 | 6.82 | 0.50 |
| medial_lemniscus | 0 | 0 | 0 | 0 | 0 | 2.33 | 1.00 |
| inferior_cerebellar_peduncle | 0 | 0 | 0 | 0 | 0 | 0 | 1.00 |
| superior_cerebellar_peduncle | 0 | 0 | 0 | 0 | 0 | 0 | 1.00 |
| cerebral_peduncle | 0 | 0 | 0.76 | 0 | 0 | 11.57 | 0.50 |
| ant_limb_int_cap | 0 | 0 | 25 | 0 | 0 | 8.60 | 0.55 |
| post_limb_int_cap | 0 | 0 | 7.55 | 0 | 0 | 5.79 | 1.00 |
| retrolent_int_cap | 0 | 0 | 7.72 | 0 | 0 | 9.81 | 1.00 |
| anterior_corona_radiata | 0 | 0 | 8.21 | 0 | 0 | 7.71 | 0.58 |
| superior_corona_radiata | 0 | 0 | 8.44 | 0 | 0 | 8.15 | 0.13 |
| posterior_corona_radiata | 0 | 0 | 9.87 | 0 | 0 | 4.42 | 0.25 |
| posterior_thalamic_radiation | 0 | 0 | 6.07 | 0 | 0 | 10.47 | 1.00 |
| sagittal | 0 | 0 | 1.04 | 0 | 0 | 19.58 | 0.22 |
| external_capsule | 0 | 0 | 16.76 | 0 | 0 | 13.03 | 0.79 |
| cingulum_cingulate_gyrus | 0 | 0 | 16.02 | 0 | 0 | 4.76 | 1.00 |
| cingulum_hippocampus | 0 | 0 | 0 | 0 | 0 | 15.03 | 0.50 |
| fornix_stria_terminalis | 0 | 0 | 5.44 | 0 | 0 | 16.79 | 0.63 |
| sup_long_fasc | 0 | 0 | 4.17 | 0 | 0 | 2.79 | 0.69 |
| sup_fronto_occipit_fasc | 0 | 0 | 18.18 | 0 | 0 | 3.39 | 0.13 |
| uncinate_fasciculus | 0 | 0 | 0 | 0 | 0 | 51.06 | 0.25 |
| tapetum | 0 | 0 | 14.71 | 0 | 0 | 7.79 | 1.00 |

Max – Maximum; Min – Minimum; n.s. – not significant.

^a^ P value for Sign tests comparing the left- and right-hemisphere lesion volumes based on quantitative GM and WM analysis. The values being compared reflect the proportion of voxels in each AAL and JHU atlas area that are included in the lesion. Minimum, median and maximum values represent the distribution across all lesions (N=40) of the proportion of voxels in each AAL and JHU atlas area that are included in a lesion.

**Table S3 –** Topography analysis comparing OCD vs. control lesions

| **OCD vs. Controls (N=40 vs. 608)** | | | | | | | | | | | |
| --- | --- | --- | --- | --- | --- | --- | --- | --- | --- | --- | --- |
| **AAL Brain Area** | | **ß±SE** | **P value** | **AAL Brain Area** | | **ß±SE** | **P value** | **JHU Brain Area** | | **ß±SE** | **P value** |
| **Frontal** | precentral_l | -0.41±0.39 | 0.30 | **Parietal** | postcentral_l | -0.25±0.30 | 0.40 | **White Matter** | middle_cerebellar_peduncle | 0.01±0.01 | 0.59 |
|  | precentral_r | -0.14±0.29 | 0.63 |  | postcentral_r | -0.29±0.25 | 0.24 |  | pontine_crossing_tract | 0.13±0.09 | 0.17 |
|  | frontal_sup_2_l | 0.40±0.39 | 0.30 |  | parietal_sup_l | -0.24±0.32 | 0.46 |  | genu_of_corpus_callosum | 1.32±0.85 | 0.12 |
|  | frontal_sup_2_r | 0.22±0.35 | 0.52 |  | parietal_sup_r | -0.18±0.18 | 0.33 |  | body_of_corpus_callosum | -0.16±0.60 | 0.79 |
|  | frontal_mid_2_l | -0.04±0.34 | 0.91 |  | parietal_inf_l | -0.42±0.45 | 0.35 |  | splenium_of_corpus_callosum | -0.49±0.49 | 0.31 |
|  | frontal_mid_2_r | 0.07±0.35 | 0.84 |  | parietal_inf_r | -0.42±0.39 | 0.29 |  | fornix | 0.43±0.74 | 0.56 |
|  | frontal_inf_oper_l | -0.27±0.68 | 0.69 |  | supramarginal_l | -0.76±0.58 | 0.19 |  | corticospinal_tract_r | 0.12±0.17 | 0.48 |
|  | frontal_inf_oper_r | 0.09±0.74 | 0.90 |  | supramarginal_r | -0.41±0.51 | 0.42 |  | corticospinal_tract_l | -0.10±0.13 | 0.45 |
|  | frontal_inf_tri_l | 0.07±0.52 | 0.89 |  | angular_l | -0.61±0.49 | 0.22 |  | medial_lemniscus_r | 0.05±0.03 | 0.13 |
|  | frontal_inf_tri_r | 0.44±0.69 | 0.52 |  | angular_r | -0.49±0.48 | 0.31 |  | medial_lemniscus_l | -0.01±0.02 | 0.70 |
|  | frontal_inf_orb_2_l | 0.71±0.47 | 0.13 |  | precuneus_l | -0.19±0.25 | 0.45 |  | inferior_cerebellar_peduncle_r | -0.01±0.03 | 0.61 |
|  | frontal_inf_orb_2_r | 0.78±0.51 | 0.13 |  | precuneus_r | -0.24±0.30 | 0.41 |  | inferior_cerebellar_peduncle_l | NA | NA |
|  | rolandic_oper_l | -0.46±0.81 | 0.57 |  | paracentral_lobule_l | -0.01±0.28 | 0.97 |  | superior_cerebellar_peduncle_r | 0.01±0.04 | 0.89 |
|  | rolandic_oper_r | -0.22±0.88 | 0.80 |  | paracentral_lobule_r | -0.16±0.15 | 0.30 |  | superior_cerebellar_peduncle_l | 0.03±0.15 | 0.86 |
|  | supp_motor_area_l | 0.40±0.33 | 0.23 | **Insula and Temporal** | insula_l | -0.27±0.76 | 0.72 |  | cerebral_peduncle_r | 0.04±0.46 | 0.93 |
|  | supp_motor_area_r | 0.07±0.29 | 0.82 |  | insula_r | 0.46±0.82 | 0.57 |  | cerebral_peduncle_l | -0.25±0.61 | 0.68 |
|  | olfactory_l | 0.32±0.62 | 0.61 |  | hippocampus_l | -0.95±0.73 | 0.19 |  | ant_limb_int_cap_r | 0.87±0.90 | 0.33 |
|  | olfactory_r | 0.41±0.61 | 0.50 |  | hippocampus_r | 0.61±0.70 | 0.38 |  | ant_limb_int_cap_l | 1.26±0.86 | 0.14 |
|  | frontal_sup_medial_l | 0.43±0.40 | 0.28 |  | parahippocampal_l | -0.26±0.46 | 0.57 |  | post_limb_int_cap_r | -0.20±1.02 | 0.85 |
|  | frontal_sup_medial_r | 0.63±0.40 | 0.12 |  | parahippocampal_r | 0.62±0.46 | 0.17 |  | post_limb_int_cap_l | -0.13±0.91 | 0.89 |
|  | frontal_med_orb_l | 0.56±0.32 | 0.08 |  | amygdala_l | -1.04±0.89 | 0.25 |  | retrolent_int_cap_r | -0.66±1.25 | 0.60 |
|  | frontal_med_orb_r | 0.50±0.44 | 0.25 |  | amygdala_r | 1.17±0.99 | 0.24 |  | retrolent_int_cap_l | -1.33±1.04 | 0.20 |
|  | **rectus_l** | **1.25±0.28** | **<0.0001** |  | heschl_l | -0.83±0.95 | 0.39 |  | anterior_corona_radiata_r | 0.98±0.84 | 0.25 |
|  | **rectus_r** | **1.15±0.34** | **0.0007** |  | heschl_r | -0.60±1.15 | 0.60 |  | anterior_corona_radiata_l | 0.89±0.83 | 0.29 |
|  | **ofcmed_l** | **1.50±0.30** | **<0.0001** |  | temporal_sup_l | -0.61±0.69 | 0.38 |  | superior_corona_radiata_r | -0.55±1.00 | 0.58 |
|  | **ofcmed_r** | **1.13±0.31** | **0.0003** |  | temporal_sup_r | 0.11±0.74 | 0.88 |  | superior_corona_radiata_l | -0.88±0.93 | 0.35 |
|  | ofcant_l | 0.33±0.17 | 0.05 |  | temporal_pole_sup_l | -0.27±0.50 | 0.60 |  | posterior_corona_radiata_r | -1.16±1.07 | 0.28 |
|  | **ofcant_r** | **0.50±0.17** | **0.004** |  | **temporal_pole_sup_r** | **1.29±0.45** | **0.005** |  | posterior_corona_radiata_l | -1.31±0.99 | 0.19 |
|  | ofcpost_l | 0.48±0.38 | 0.21 |  | temporal_mid_l | -0.52±0.50 | 0.30 |  | posterior_thalamic_radiation_r | -0.10±0.97 | 0.92 |
|  | ofcpost_r | 0.86±0.37 | 0.02 |  | temporal_mid_r | 0.61±0.47 | 0.20 |  | posterior_thalamic_radiation_l | -0.92±0.87 | 0.29 |
|  | ofclat_l | 0.05±0.20 | 0.81 |  | temporal_pole_mid_l | -0.15±0.22 | 0.50 |  | sagittal_stratum_r | 0.51±0.78 | 0.51 |
|  | ofclat_r | 0.35±0.18 | 0.06 |  | **temporal_pole_mid_r** | **0.76±0.26** | **0.004** |  | sagittal_l | -1.40±0.93 | 0.13 |
| **Cingulum** | cingulate_ant_l | 0.88±0.59 | 0.14 |  | temporal_inf_l | -0.25±0.30 | 0.41 |  | external_capsule_r | 0.37±1.06 | 0.72 |
|  | cingulate_ant_r | 0.89±0.72 | 0.22 |  | temporal_inf_r | 0.28±0.26 | 0.28 |  | external_capsule_l | -0.02±1.01 | 0.98 |
|  | cingulate_mid_l | -0.16±0.49 | 0.74 | **Cerebellum** | cerebelum_crus1_l | 0.01±0.01 | 0.47 |  | cingulum_cingulate_gyrus_r | -0.21±0.70 | 0.77 |
|  | cingulate_mid_r | -0.24±0.49 | 0.63 |  | cerebelum_crus1_r | -0.01±0.03 | 0.66 |  | cingulum_cingulate_gyrus_l | 0.00±0.75 | 1.00 |
|  | cingulate_post_l | -0.37±0.47 | 0.44 |  | cerebelum_crus2_l | 0.06±0.06 | 0.31 |  | cingulum_hippocampus_r | 0.42±0.53 | 0.43 |
|  | cingulate_post_r | -0.39±0.68 | 0.56 |  | cerebelum_crus2_r | -0.03±0.09 | 0.74 |  | cingulum_hippocampus_l | -0.23±0.73 | 0.75 |
| **Subcortical Grey Matter** | caudate_l | 1.02±0.71 | 0.15 |  | cerebelum_3_l | 0.04±0.23 | 0.85 |  | fornix_stria_terminalis_r | 0.31±1.08 | 0.78 |
|  | caudate_r | 0.91±0.77 | 0.24 |  | cerebelum_3_r | 0.04±0.08 | 0.63 |  | fornix_stria_terminalis_l | -0.79±0.92 | 0.39 |
|  | putamen_l | 0.83±0.96 | 0.38 |  | cerebelum_4_5_l | 0.05±0.18 | 0.76 |  | sup_long_fasc_r | -0.75±0.83 | 0.37 |
|  | putamen_r | 0.42±1.05 | 0.69 |  | cerebelum_4_5_r | 0.06±0.08 | 0.42 |  | sup_long_fasc_l | -1.34±0.82 | 0.10 |
|  | pallidum_l | 1.93±1.08 | 0.08 |  | cerebelum_6_l | 0.05±0.06 | 0.43 |  | sup_fronto_occipit_fasc_r | -0.86±2.08 | 0.68 |
|  | pallidum_r | 0.65±1.26 | 0.61 |  | cerebelum_6_r | -0.02±0.06 | 0.68 |  | sup_fronto_occipit_fasc_l | 0.15±1.59 | 0.93 |
|  | thalamus_l | 0.08±0.56 | 0.88 |  | cerebelum_7b_l | 0.15±0.07 | 0.03 |  | uncinate_fasciculus_r | 1.36±1.28 | 0.29 |
|  | thalamus_r | -0.04±0.75 | 0.95 |  | cerebelum_7b_r | 0.00±0.02 | 0.77 |  | uncinate_fasciculus_l | -1.79±1.28 | 0.16 |
| **Occipital** | calcarine_l | -0.11±0.20 | 0.56 |  | cerebelum_8_l | 0.01±0.06 | 0.90 |  | tapetum_r | -0.51±1.17 | 0.66 |
|  | calcarine_r | 0.00±0.35 | 0.99 |  | cerebelum_8_r | 0.00±0.01 | 0.81 |  | tapetum_l | -0.45±1.01 | 0.65 |
|  | cuneus_l | -0.23±0.28 | 0.41 |  | cerebelum_9_l | -0.01±0.02 | 0.67 |  | | | |
|  | cuneus_r | -0.17±0.29 | 0.54 |  | cerebelum_9_r | NA | NA |  |  |  |  |
|  | lingual_l | -0.01±0.29 | 0.98 |  | cerebelum_10_l | NA | NA |  |  |  |  |
|  | lingual_r | 0.17±0.28 | 0.55 |  | cerebelum_10_r | -0.01±0.04 | 0.78 |  |  |  |  |
|  | occipital_sup_l | -0.32±0.38 | 0.41 |  | vermis_1_2 | 0.04±0.13 | 0.75 |  |  |  |  |
|  | occipital_sup_r | -0.24±0.33 | 0.47 |  | vermis_3 | 0.03±0.17 | 0.86 |  |  |  |  |
|  | occipital_mid_l | -0.31±0.27 | 0.26 |  | vermis_4_5 | -0.01±0.05 | 0.80 |  |  |  |  |
|  | occipital_mid_r | 0.04±0.30 | 0.90 |  | vermis_6 | -0.08±0.16 | 0.61 |  |  |  |  |
|  | occipital_inf_l | -0.12±0.32 | 0.72 |  | vermis_7 | -0.07±0.14 | 0.61 |  |  |  |  |
|  | occipital_inf_r | 0.28±0.22 | 0.20 |  | vermis_8 | -0.01±0.05 | 0.81 |  |  |  |  |
|  | fusiform_l | -0.21±0.29 | 0.48 |  | vermis_9 | 0.00±0.01 | 0.81 |  |  |  |  |
|  | fusiform_r | 0.33±0.28 | 0.25 |  | vermis_10 | NA | NA |  |  |  |  |

ß – Beta coefficient from the general linear model; _l – left; _r – right; AAL – Automated Anatomical Labeling atlas; JUH – John Hopkins University atlas; NA – Not Applicable; OCD – Obsessive-Compulsive Disorder; SE – Standard Error from the general linear model.

Comparison between lesional OCD and control lesion cohort was conducted using general linear models for each region of interest of grey and white matter atlases, controlling for lesion size and lesion etiology (vascular vs. non-vascular). Gabbay et al. 2003 and Tonkonogy et al. 1989 classified as non-vascular lesions, despite no specific etiology has been reported in both case reports.

* Regions and values highlighted in bold reflect significant defined using a False Discovery Rate (FDR) of 0.1, according to Benjamini-Hochberg^8^.

**Table S4 –** Spatial correlations between original analyses with those resulting from the reliability and robustness analyses

| **Reliability & Robustness**  **of OCD vs. Controls Map** | | **Pearson’s Correlation** | |
| --- | --- | --- | --- |
|  |  | **Topography** | **Connectivity** |
| Inter-rater (OCD=40 vs. Controls=608) | | 0.94 | 0.98 |
| Intra-rater (OCD=40 vs. Controls=608) | | 0.95 | 1.00 |
| Wilcoxon Rank-Sum Test (OCD=40 vs. Controls=608) | | 0.76 | N.A. |
| HCP Connectome (OCD=40 vs. Controls=608) | | N.A. | 0.85 |
| Auto-connectivity | No OFC (OCD=32 vs. Controls=586)^a^  No OFC (OCD=32 vs. Controls=608)^b^ | N.A.  N.A. | 0.91  0.92 |
|  | No BG (OCD=35 vs. Controls=545)^a^  No BG (OCD=35 vs. Controls=608)^b^ | N.A.  N.A. | 0.98  0.99 |
|  | No OFC or BG (OCD=31 vs. Controls=539)^a^  No OFC or BG (OCD=31 vs. Controls=608)^b^ | N.A.  N.A. | 0.91  0.92 |
| Half 1 (OCD=20 vs. Controls=304) | | 0.87 | 0.95 |
| Half 2 (OCD=20 vs. Controls=304) | | 0.82 | 0.86 |
| Full DSM 5 Criteria (OCD=30 vs. Controls=608) | | 0.98 | 0.98 |
| Y-BOCS ≥14^c^ (OCD=14 vs. Controls=608) | | 0.89 | 0.79 |
| No Meds at Onset (OCD=33 vs. Controls=608) | | 0.99 | 0.99 |
| No NP risk^d^ (OCD=28 vs. Controls=608) | | 0.95 | 0.96 |
| Early Latency^e^ (OCD=20 vs. Controls=608) | | 0.88 | 0.96 |
| No TBI or Infec. (OCD=33 vs. Controls=608) | | 0.87 | 0.96 |
| CQA≥4 (OCD=22 vs. Controls=608) | | 0.94 | 0.97 |
| BLDA≥4 (OCD=33 vs. Controls=608) | | 0.99 | 0.99 |
| Only MRI (OCD=29 vs. Controls=608) | | 0.98 | 0.99 |

BLDA – Brain Lesion Documentation Assessment; CQA – Clinical Quality Assessment; DSM – Diagnostic and Statistical Manual of Mental Disorders; Infec. – Infectious etiology; MRI – Magnetic Resonance Imaging; NP – Neuropsychiatric Disorder; OCD – Obsessive Compulsive Disorder; TBI – Traumatic brain injury; Y-BOCS – Yale-Brown Obsessive Compulsive Scale.

^a^ Excluding lesions in both cohorts.

^b^ Excluding lesions only in lesional OCD cohort.

^c^ Y-BOCS≥14 was selected to define moderate to severe OCD according to current literature^9^.

^d^ No neuropsychiatric disorder risk was defined by excluding cases with personal or family history of neuropsychiatric disorders.

^e^ Early latency was defined in accordance with the temporal association between lesion onset and emergence of OCD-like symptoms, splitting the sample in two subgroups according to the median time between the event and the emergence of symptoms (302.5). Early latency sub-group consisted in cases with documented temporal association of 240 days or less (N=20).

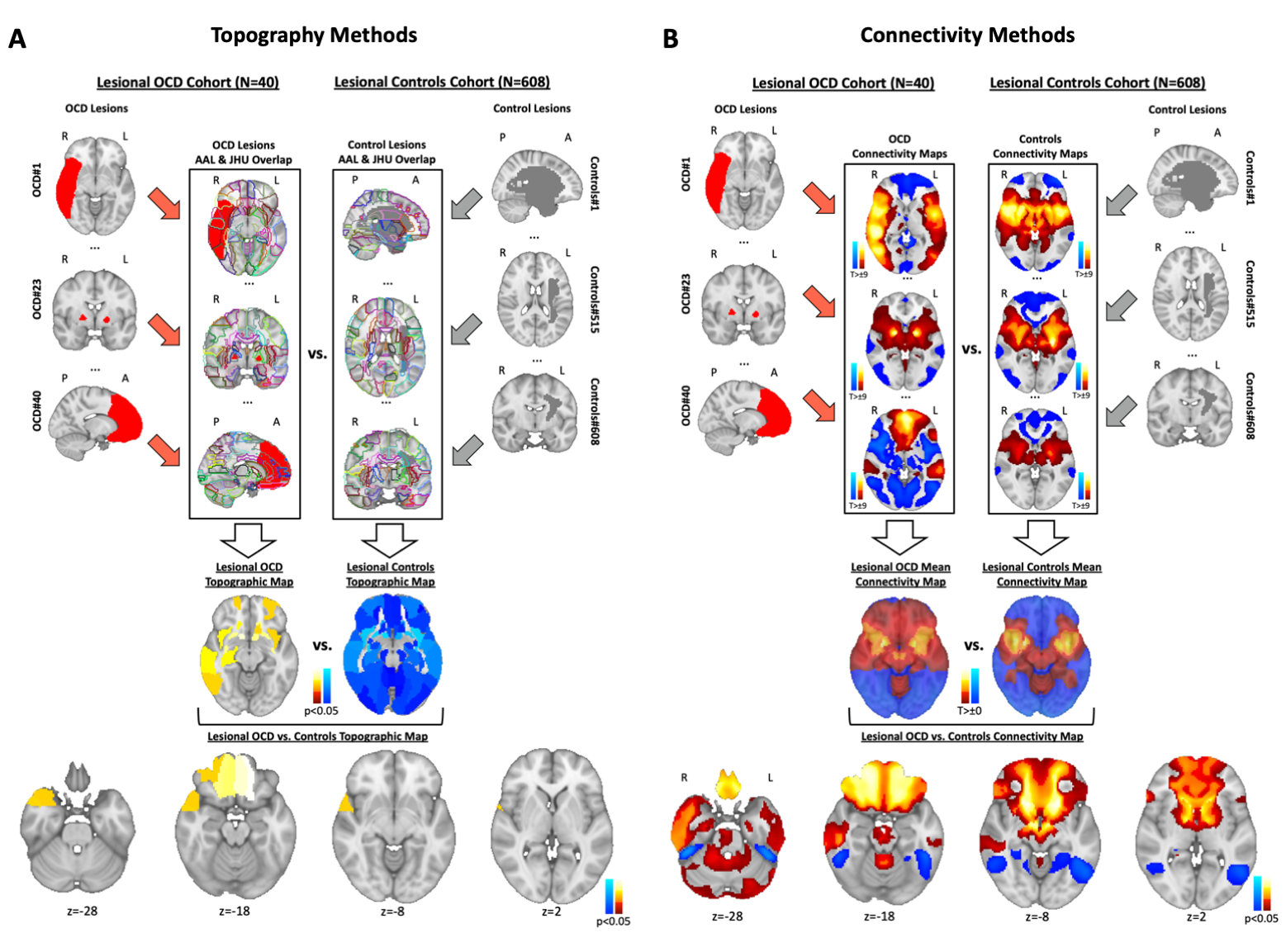
**SUPPLEMENTARY FIGURES**

**Figure S1. Computing topography and connectivity maps.** Lesion locations associated with OCD-like symptoms (N=40), or control lesions (N=608) were mapped onto the MNI standard brain. **A.** For topography analyses, we compared the proportion of affected voxels in each area of a grey (AAL) and white (JHU) matter atlases, between lesional OCD and controls cohorts. Initially, for each area of the atlases, we performed a one sample sign test using the proportion of affected voxels, separately for OCD lesions, computing the “Lesional OCD Topography Map”, and for control lesions. Warm and cold colors represent areas that are more intersected by OCD or by control lesions, respectively, in the respective maps. The Lesional OCD vs. Controls Topography Map was computed using a general linear model for each of these areas, controlling for lesion size and lesion etiology, and statistical significance was defined according to Benjamini-Hochberg^8^, assuming a false discovery rate (FDR) of 0.1, to control for multiple comparisons. **B.** For the connectivity analyses, based on a large normative resting-state functional connectivity database, we found brain regions that were functionally connected to each lesion location, to compare the functional connectivity patterns obtained from OCD and control lesion locations. Initially, all OCD connectivity maps were averaged to obtain the Lesional OCD Mean Connectivity Map, and an equivalent computation for control lesions. In both the Lesional OCD and control mean connectivity maps, warm and cold colors represent areas that are respectively more positively and negatively connected to OCD or to control lesions. The Lesional OCD vs. Controls Connectivity Map was computed using a voxel-wise permutation-based two-sample t-test performed within FSL PALM (5000 permutations), displayed at an FWE-corrected level of p<0.05. In both OCD vs. Controls maps, warm and cold colors represent areas that are more or less intersected or connected to OCD lesions when compared to controls, respectively.

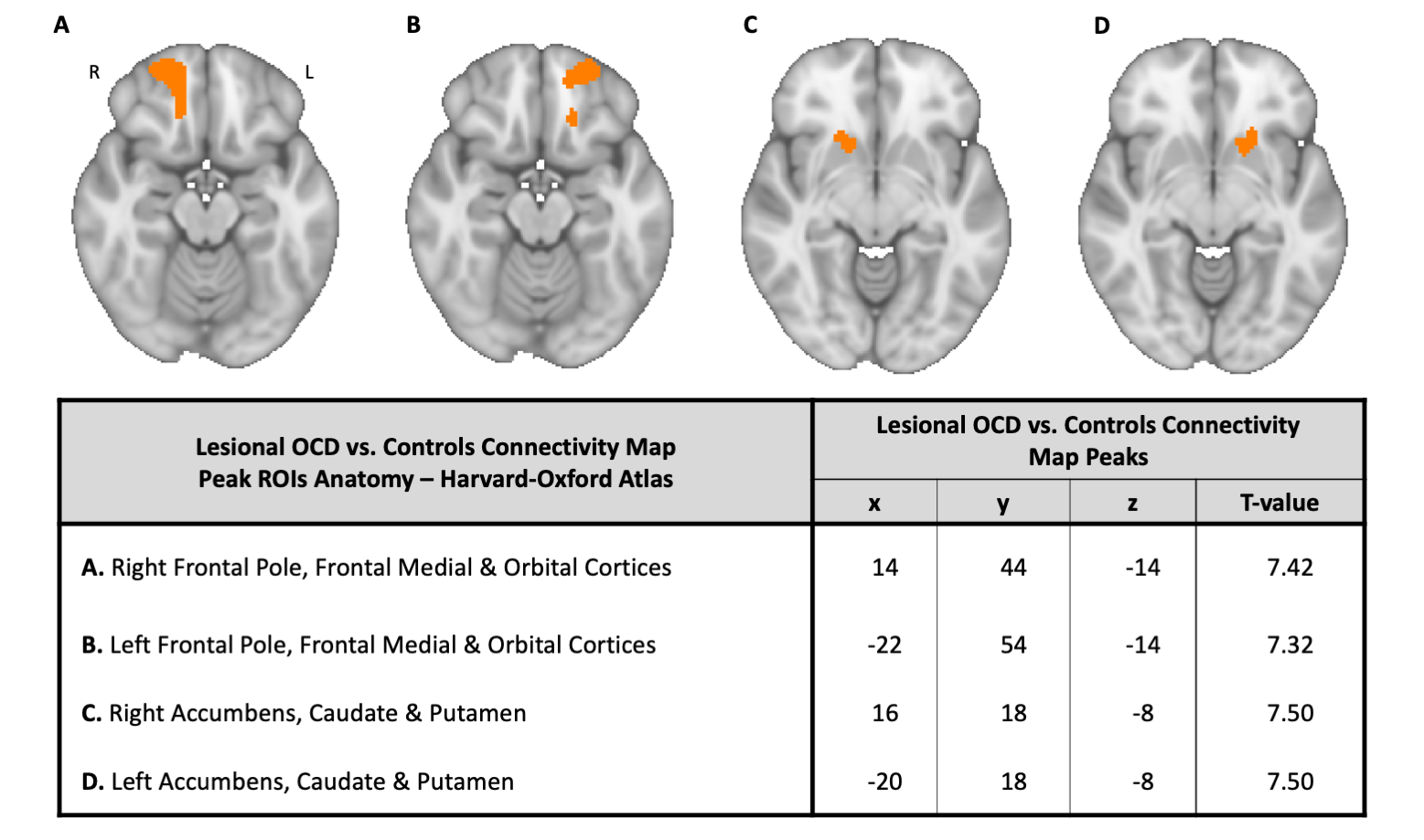
**Figure S2. Most specific and significant lesional OCD functional connectivity regions.** Peak regions (**A**, **B**, **C** and **D**) in Lesional OCD vs. Controls Connectivity Map and respective peak coordinates. Peak regions of this map were considered if p_FWE_<4.1x10^-5^ and a minimum volume of 200mm^3^.

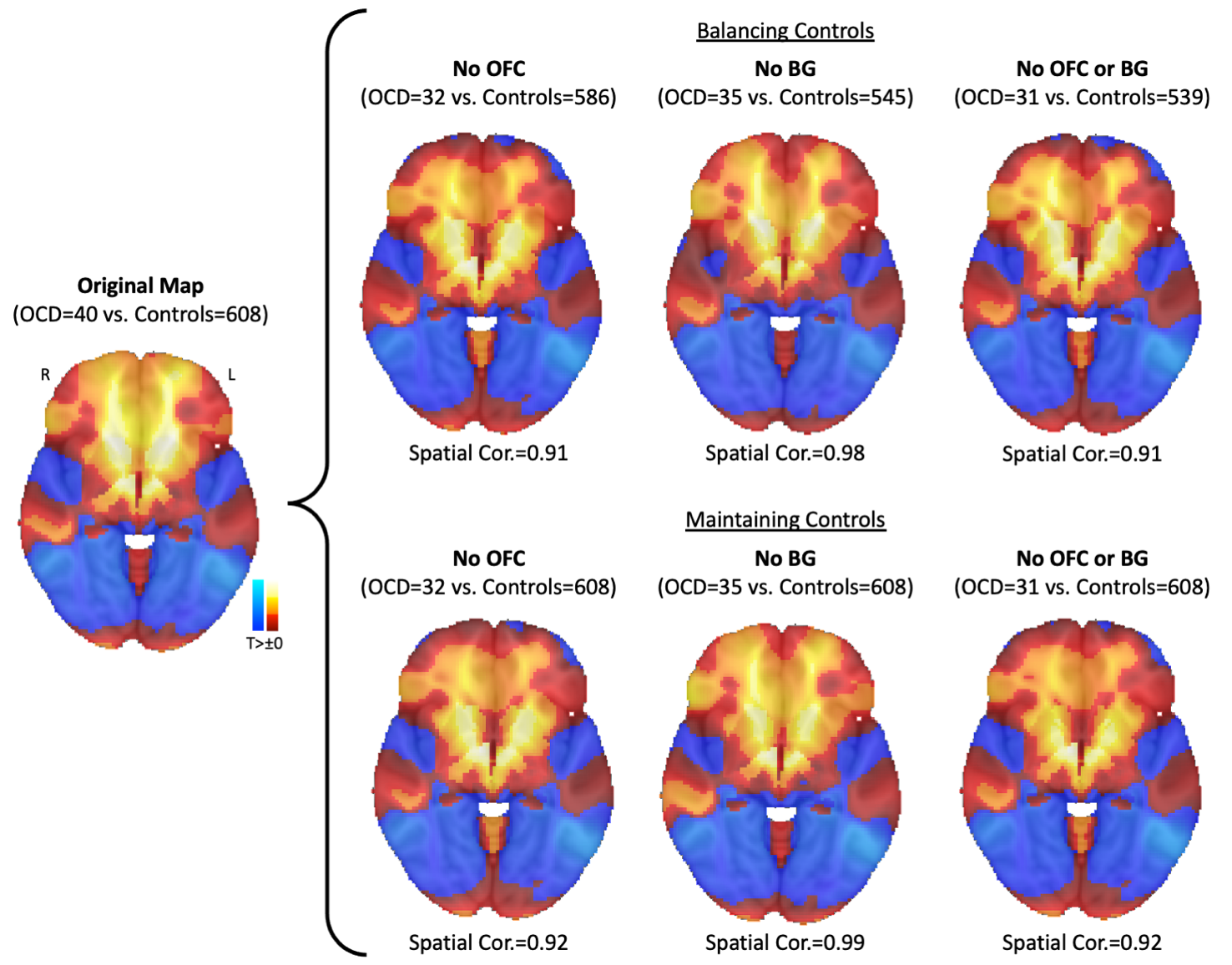
**Figure S3. Lesional OCD networks extend beyond lesion location.** Lesional OCD vs. Controls Connectivity pattern extend beyond those explained by lesion location alone, since Lesional OCD vs. Controls Connectivity Maps were similar to the original even when excluding lesions located within peak connectivity regions in the OFC and BG (Figure S2). Similar results were obtained irrespective of excluding lesions with these characteristics in both samples (**Balancing Controls**) or only in lesional OCD cohort (**Maintaining Controls**). In all maps, warm and cold colors represent areas that are more or less connected to OCD lesions when compared to controls, respectively.

BG – Basal Ganglia; Cor. – Correlation FEW – family wise error; MNI – Montreal Neurological Institute; OCD – obsessive compulsive disorder; OFC – orbitofrontal cortex.

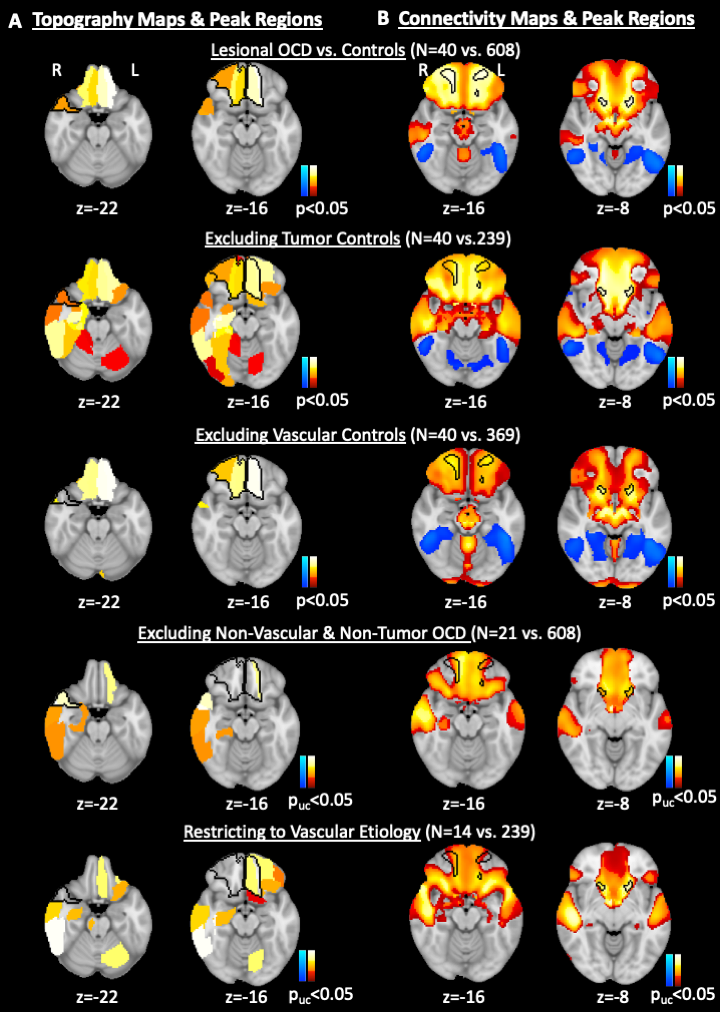
**Figure S4. Lesion etiology may confound topography of OCD lesions but not their connectivity.** Despite the original analyses being computed controlling for lesion etiology, we further explored the impact of lesion etiology on our results. **A.** The regions identified in the Lesional OCD vs. Controls Topography map were different when excluding tumor or vascular controls, when excluding non-vascular and non-tumor OCD lesions and when restricting both samples only to vascular etiology. The original topography regions encompassed right anterior and medial OFC, and rectus gyrus, left medial OFC and rectus gyrus, and right middle and superior temporal pole (**Black Outline** in panel **A** maps). **B.** On the other hand, peak regions in the original Lesional OCD vs. Controls Connectivity Map were consistently part of new connectivity maps excluding tumor and/or vascular OCD and/or control lesions. Peak connectivity regions included right frontal pole, medial frontal cortex, and OFC, left frontal pole and OFC, and right and left accumbens, caudate and putamen (**Black Outline** in panel **B** maps). The maps in the first three rows of both panels A and B were corrected for multiple comparisons according to Benjamini-Hochberg^8^ and displayed at an FDR-corrected level of p<0.05 for topography maps or using threshold free cluster enhancement and displayed at an FWE-corrected level of p<0.05 for connectivity maps. In the last two rows of both panels A and B we show uncorrected p-values since approximately 50% or more of our sample of interest, i.e., lesional OCD cases, was excluded. In all maps, warm and cold colors represent areas that are more or less insulted (panel A) or connected (panel B) to OCD lesions when compared to controls, respectively.

FDR – false discovery rate; FEW – family wise error; MNI – Montreal Neurological Institute; OCD – obsessive compulsive disorder; OFC – orbitofrontal cortex; uc – uncorrected.

**SUPPLEMENTARY MATERIAL REFERENCES**
